# In Silico Modeling of Transcatheter Heart Valve Oversizing and Ellipticity, Part II: Effects on Leaflet Mechanics, Hemodynamics, and Stent Deflection Contributing to Thrombogenic Risk and Structural Degeneration

**DOI:** 10.1101/2025.09.25.25336635

**Authors:** Sam Boxwell, Rachel M.E. Cahalane, Dylan Armfield, William Hickey, Scott Cook, Patricia Kelly, Philip Cardiff, Laoise McNamara

## Abstract

**Background and Objectives:** Transcatheter aortic valve implantation (TAVI) is the leading treatment for aortic stenosis. Self-expanding transcatheter heart valves (THVs) are oversized to prevent paravalvular leakage and then deployed over the diseased native valve. However, this can result in incomplete expansion and elliptical deployment, which may influence thrombogenic risk and structural degeneration, although this is not fully understood.

**Methods:** In this study, we utilized a validated *in silico* framework to assess the impact of THV oversizing and ellipticity on leaflet mechanics, hemodynamic shear stress and stent deformation, which are indicators of structural degeneration and thrombogenicity. We simulated self-expansion of a deformable THV stent within an idealized aortic annulus, applied pulsatile loading conditions representative of the cardiac cycle and then evaluated post-deployment frame deformation, leaflet mechanics, hemodynamics and stent fatigue.

**Results:** We predicted ‘stent-frame decoupling’ of the supra-annular THV, with increased expansion and circularity at the functional valve level compared to the inflow. Maximal oversizing of the THV reduced valve expansion at the supra-annular valve level (< 90% expansion), which increased leaflet coaptation and pinwheeling, but reduced peak leaflet stresses and stent deflection. Maximal oversizing also altered hemodynamics, causing early mainstream flow separation, which increased leaflet oscillatory shear and viscous shear stress downstream of the THV, potentially increasing thrombogenic risk and promoting tissue degeneration. THV ellipticity induced heterogenous stent deflections, leading to variable leaflet stress distributions and coaptation mismatch.

**Conclusion:** We propose that flexible THV stents may mitigate adverse effects of elliptical deployment and emphasize the importance of assessing THV expansion through fluoroscopy and considering post-TAVI balloon-dilatation to increase expansion and improve long-term functional valve performance.

## 1 Introduction

Aortic stenosis is characterized by progressive fibro-calcific remodeling of the aortic valve, impairing leaflet motion and inhibiting normal valvular function. It is currently the most prevalent valvular heart disease in developed countries, and incidence is expected to increase significantly with an aging population – positioning it as the ‘next cardiac epidemic’ [1]. Traditionally, treatment of this condition involved open-heart surgical aortic valve replacement (SAVR). However, the minimally-invasive transcatheter aortic valve implantation (TAVI) has become the primary treatment for aortic stenosis in older patients (87% in 65-80 years), and up to 50% of younger cohorts (> 65) [2–4]. This procedure involves percutaneous delivery of a transcatheter heart valve (THV), which is deployed by balloon- or self-expansion over the diseased native valve. The final deployment morphology of THVs is dependent on the interaction of the device with the native anatomy, which may vary greatly between patients due to differences in the aortic root and diseased aortic valve structures. Unlike balloon-expandable devices, which reshape the native anatomy through plastic deformation of the metallic stent, self-expanding THVs rely on the super-elastic properties of nitinol for expansion and thereby conform to the aortic annulus geometry. These devices typically exhibit lower radial force and require excessive oversizing (5-16%) relative to the native annulus perimeter to prevent paravalvular leakage, when compared to balloon-expandable devices (up to 5%) [5, 6]. Furthermore, calcium deposits, located on the diseased native valve, can exacerbate asymmetric valve distortion and prevent complete stent deployment. Together, these factors lead to varied expansion and elliptical frame deformation of self-expanding THVs, which alters device kinematics and hemodynamics [7–9].

The environment resulting from varied THV expansion and ellipticity may induce calcification, leaflet rupture, thrombus formation and structural valve degeneration (SVD) [8, 10–12], leading to early device failure [8, 13]. Leaflet mechanics are altered by excessive oversizing and elliptical deployment of THVs, which may increase tensile, compressive and flexural stresses in regions of bioprosthetic tissue, thereby damaging collagen fibrils and exposing potential calcium-binding sites [10, 11, 14]. In addition, fluid-induced wall shear stress (WSS) patterns may be impacted by THV deployment [12, 15, 16] and have been correlated with spatial distribution of calcification across the tissue leaflets of explanted SAVRs [12]. Platelets and macrophages have been observed on bioprosthetic leaflet surfaces [17, 18] and are likely influenced by turbulent flow and viscous shear stresses, which may lead to platelet activation and thrombus formation [19]. Activated platelets may travel downstream and pose a risk of stroke, or be directed within the annulus, where low-flow conditions promote adhesion and thrombosis [16, 20]. Such flow conditions may also influence other cells involved in calcification and thrombosis of bioprosthetic tissue, but this remains poorly understood. Although self-expanding THVs have demonstrated durability up to 6-years [21], the long-term performance of the THV stent will become increasingly important as TAVI extends to younger patient cohorts. These devices experience complex loading in vivo, due to the force of the stent against the annulus [22] and leaflet loading [23], which can vary depending on expansion and deployment configurations. As THVs will be required to sustain performance for longer durations, there is a need to further understand the influence of deployment on stent deformation and fatigue.

During device development, THV performance can be assessed through radial force, high-speed dynamic imaging, particle image velocimetry [24] and accelerated wear testing (ISO 5840-3), with leaflet and stent testing to 200 and 400 million cycles (equivalent to -5 and -10 years *in vivo*) [25]. Numerical simulation can complement *in vitro* testing and provide valuable predictions of tissue strain, shear stress and stagnation fields, to provide a more comprehensive insight into THV biomechanics, hemodynamics and long-term device function [23]. Previous computational studies have used computational solid mechanics, computational fluid dynamics (CFD) or fluid-structure interaction (FSI) approaches to investigate the impact of oversizing and elliptical deployment of THVs [7, 26–28]. These studies predict that elliptical deployment of THVs impairs valvular kinematics and hemodynamics, by increasing peak leaflet stress, increasing flow stasis and directing regions of high viscous shear stress and turbulent flow into the sinus of Valsalva [7, 26–28]. Even moderate oversizing of self-expanding THVs (between 2.5-9.5% by perimeter [6]) may increase leaflet coaptation, pinwheeling and particle residence time [29–31]. However, previous computational studies examining the impact of THV deployment have not accounted for interactions between the flexible stent frame and anisotropic leaflets, but assume stent rigidity during cardiac function [7, 26, 29]. Supra-annular, self-expanding THVs (e.g. ACURATE Prime – Boston Scientific, EVOLUT-Pro – Medtronic) exhibit notable stent deflections due to pulsatile loading [23, 32], which we recently accounted for in a computational assessment of THV function [15][24]. Furthermore, previous computational modeling studies have not accounted for ‘stent-frame decoupling’, which has been observed in clinical studies of supra-annular THVs [33, 34], as they assumed intra-annular deployment [7] or uniform expansion [28, 29]. This occurs when the inflow region of the THV conforms to the native annulus and exhibits reduced expansion and ellipticity, while the supra-annular valve functional region maintains a more expanded, circular configuration [34]. The application of advanced modeling approaches, which include stent-leaflet interactions, is required to study how deployment of supra-annular, self-expanding THVs may impact long-term device function.

In this paper, the second part of a comprehensive two-part computational study, we investigate how deployment conditions of a supra-annular, self-expanding THV influences biomechanical and hemodynamic indices of long-term device function. In part one, we established the credibility of our computational framework through comprehensive verification and validation according to ASME VV-40 [24]. The specific objective of part two of this study is to determine the impact of THV oversizing and ellipticity on leaflet stress, stent deformation and hemodynamics across a range of clinically-relevant deployment configurations.

## 2 Methods

### 2.1 Structural and hemodynamic simulation framework

As outlined in part one of this study, a finite element (FE) model of the ACURATE Prime XL was developed using Abaqus/Explicit (v2023, Simulia, Dassault Systémes, Providence, RI, USA), where the device model included the porcine pericardium leaflets, an inner sealing skirt, outer skirt and the nitinol stent, with relevant device geometries obtained from Boston Scientific (Galway, Ireland). Calculation verification informed the selection of the mesh density, element formulation and target-time increment [24]. The self-expanding THV stent was meshed with linear solid elements with reduced integration with a global element size of 0.15mm, whilst the leaflet, inner skirt and outer skirt were meshed with linear hexahedral incompatible mode (C3D8I) elements with second order accuracy [7] with a global element size of 0.25 mm. The complete numerical model of the ACURATE Prime XL THV is shown in Figure 1, with element and mesh discretization listed in Table 1.

**Figure 1:**
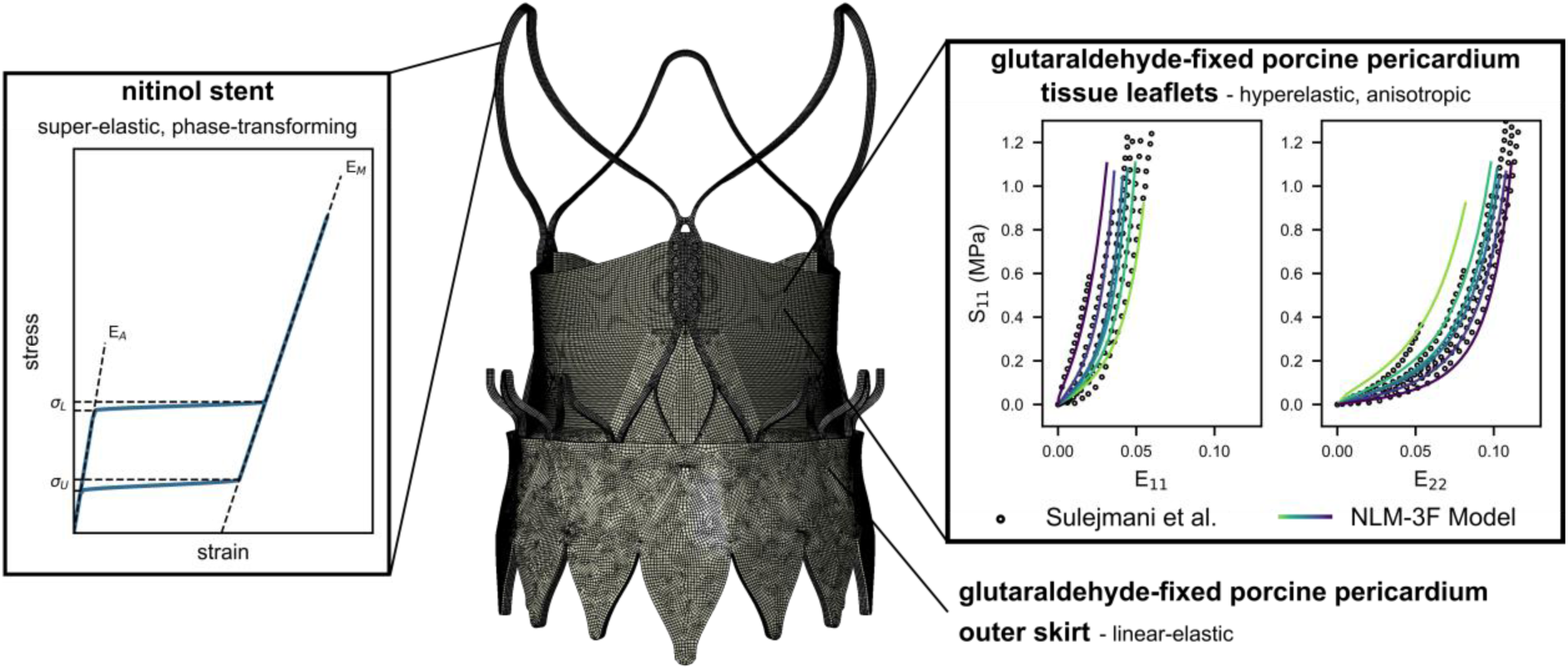
Numerical model of ACURATE Prime XL device showing device geometry, mesh density, device components (nitinol stent, porcine pericardial tissue leaflets and skirt) and their respective material models.

**Table 1:** Element details for numerical model of THV.

| Component | Element Type | Element Spacing | Element # | Thickness (mm) | # of Elements through Thickness |
| --- | --- | --- | --- | --- | --- |
| Stent | C3D8R | 0.15 | 76,668 | 0.60 | 4 |
| Leaflet and inner skirt | C3D8I w/ second order accuracy | 0.25 | 140,634 | 0.25 | 3 |
| Outer skirt | C3D8I w/ second order accuracy | 0.25 | 28,666 | 0.20 | 1 |

**Table 2:** Non-linear matrix, 3-fibre (NLM-3F) model parameters for glutaraldehyde-fixed porcine pericardium.

| C<sub>10</sub> (MPa) | C<sub>20</sub> (MPa) | C<sub>30</sub> (MPa) | k<sub>1</sub> (MPa) | k<sub>2</sub> | k<sub>3</sub> (MPa) | k<sub>4</sub> |
| --- | --- | --- | --- | --- | --- | --- |
| 0.05 | 4.13 | 4.86 | 1.00 | 65.97 | 1.32 | 58.64 |

Glutaraldehyde-fixed porcine pericardium bioprosthetic components were assumed to be anisotropic and hyperelastic, in which we used a novel constitutive material model, deemed the non-linear matrix, 3-fibre (NLM-3F) model, as previously described in part one of this study [24]. To compute the NLM-3F model parameters, a least-squares fitting approach, with respect to the second Piola–Kirchhoff stress, was employed and fit to multi-protocol stress-strain curves of glutaraldehyde-fixed porcine pericardium obtained from Sulejmani *et al.* [35], where the fitting tool is openly available on <u>Github</u>. The resulting

NLM-3F model approximations are presented in Figure 1 and Table 1. The nitinol self-expanding stent frame (Figure 1) was treated as a phase-transforming, superelastic material [15][24]. The material was assumed to have a density of 6,450 kg/m^3^ and a damping coefficient (α = 35 /s) was used to dampen oscillations in the structure.

The workflow outlined in part one of this study was used to simulate THV crimping, deployment and cardiac cycle device dynamics and hemodynamics, as presented in Figure 2. Briefly, the simulation workflow comprised the following steps:

1. *Crimping of the THV inside the catheter.* The complete device was crimped to a final outer diameter of 10 mm using 12 rigid plates which were positioned concentrically around the device.
2. *‘Top-down’ deployment of the THV device inside the aortic annulus.* The device was deployed through a two-step process which involved axial displacement of 12 upper rigid plates and subsequently, axial displacement of the 12 lower plates.
3. *Simulation of cardiac cycle device dynamics through FE modeling.* Hypertensive physiological pressure waveforms, operating at pressure waveform replicating a cardiac cycle of 70 BPM (0.86 s) with a peak trans-aortic pressure of 140 ± 3 mmHg (**Error! Reference source not found.**(D)), were applied to the aortic and ventricular surfaces of the leaflets. To reduce computational cost, we applied a variable semi-automated mass-scaling strategy with a maximum time step of 1.0 × 10^−6^, whilst ensuring that the ratio of kinetic energy to internal energy did not exceed 5% [24]. The complete FE simulation, including crimping, deployment and two cardiac cycles, was solved using 128 CPU cores (2.35 GHz AMD Rome CPUs), with wall clock times ranging from 32 - 34 hours. Following completion of the FE simulation, we extract the peak systolic device configuration for:
4. *Simulation of systolic flow using CFD modeling.* Peak systolic device dynamics obtained from the FE simulations described above were integrated into independent fluid flow simulations. We then performed a transient flow analysis in OpenFOAM using the PISO-SIMPLE (PIMPLE) algorithm, by applying a time-dependent pressure gradient. The models were solved for 3 cardiac cycles, where a dynamic time-stepping scheme was chosen to ensure the Courant-Friedrichs-Lewy (CFL) number was below 1.0 at all times.

**Figure 2:**
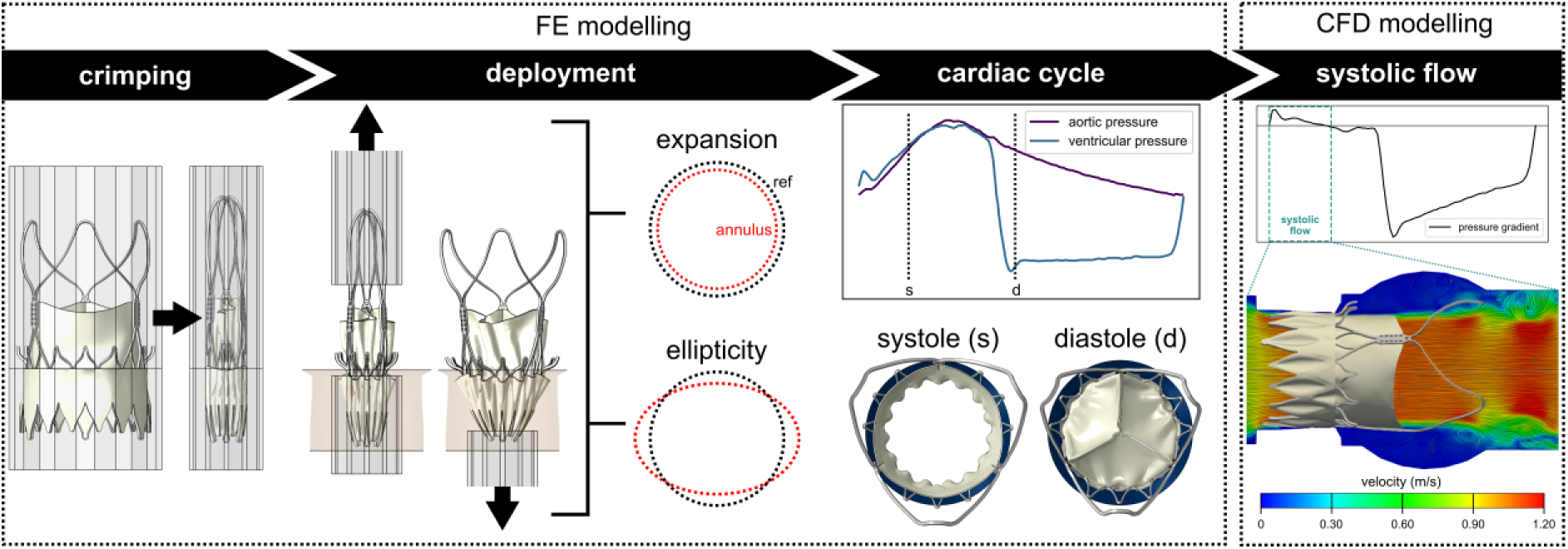
In silico structural and hemodynamic modeling framework, showing finite element simulation of THV crimping to 10mm, deployment of the device in the aortic annulus for various degrees of expansion and ellipticity, application of pulsatile cardiac cycle loading conditions and computational fluid dynamics analysis of systolic flow.

### 2.2 Frame expansion and ellipticity

To investigate the impact of device deployment, the THV was implanted within various annuli geometries, chosen to reflect clinical variability in terms of device expansion and ellipticity [5, 36]. These geometries were characterized using the oversizing and frame ellipticity indices, relative to the size of the ACURATE Prime XL (29 mm - recommended for annulus perimeter derived diameters ranging from 26.5 mm to 29 mm), defined as:

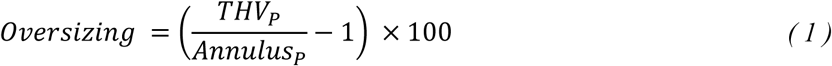

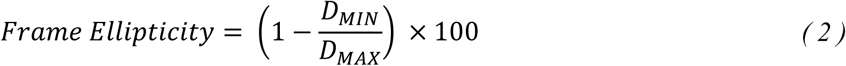

where *THV*_*P*_ and *Annulus*_*P*_ is the perimeter of the device (29 mm) and annulus respectively, and *D*_*MIN*_ and *D*_*MAX*_ are the minimum and maximum annulus diameters respectively. In part one of this study, we validated device models across various deployment configurations, which we classify as (1) ‘min-oversizing, circular’, (2) ‘max-oversizing, circular’ and (3) ‘min-oversizing, elliptical’, where min refers to minimum-oversizing (0%) and max refers to maximum-oversizing (9.4%) as indicated for the self-expanding THV. To assess the synergistic effect of both oversizing and ellipticity, in this study, we also simulate device deployment in an additional annulus, classified as; (4) ‘max-oversizing, elliptical’, where the oversizing and frame ellipticity are equal to 9.4% and 24% respectively at the annular level. Figure 3 shows the THV implanted within the deployment configurations investigated in this study, with corresponding oversizing and ellipticity indices [36, 37]. Recent clinical studies have observed ‘stent-frame decoupling’, where device expansion and ellipticity differ at the inflow, annular and supra-annular level [34]. To evaluate post-deployment frame geometry and investigate the degree of stent-frame decoupling, we compare the stent frame expansion (equation 3 below [33]) and frame ellipticity indices;

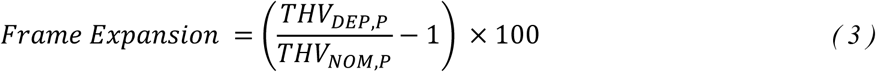

**Figure 3:**
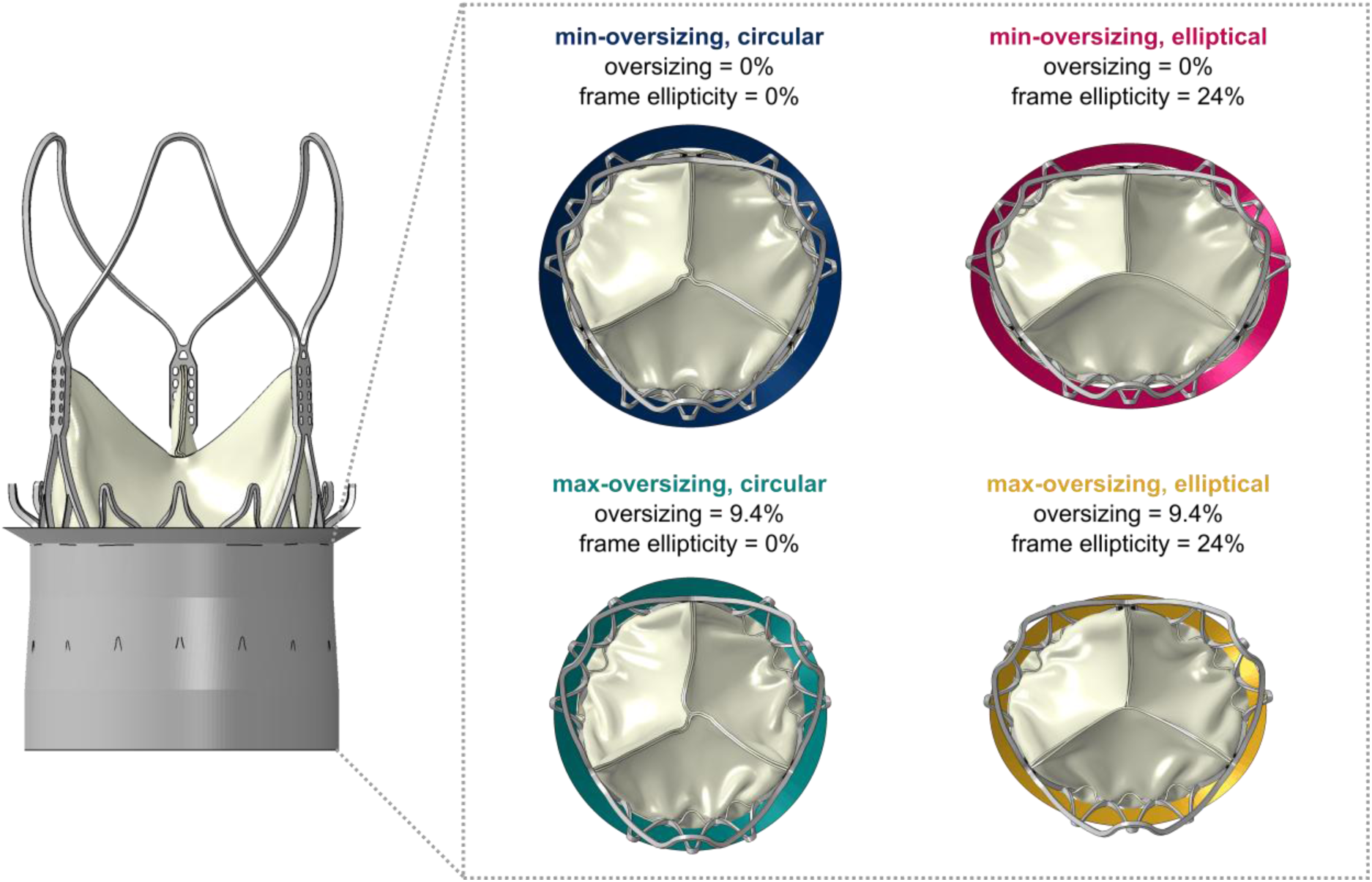
THV expansion and elliptical deployment at annular level calculated using labelled device diameter.

where *THV*_*DEP*,*P*_ and *THV*_*NOM*,*P*_ are the cross-sectional perimeters of the deployed and nominal device at each frame plane level respectively (Figure 4).

**Figure 4:**
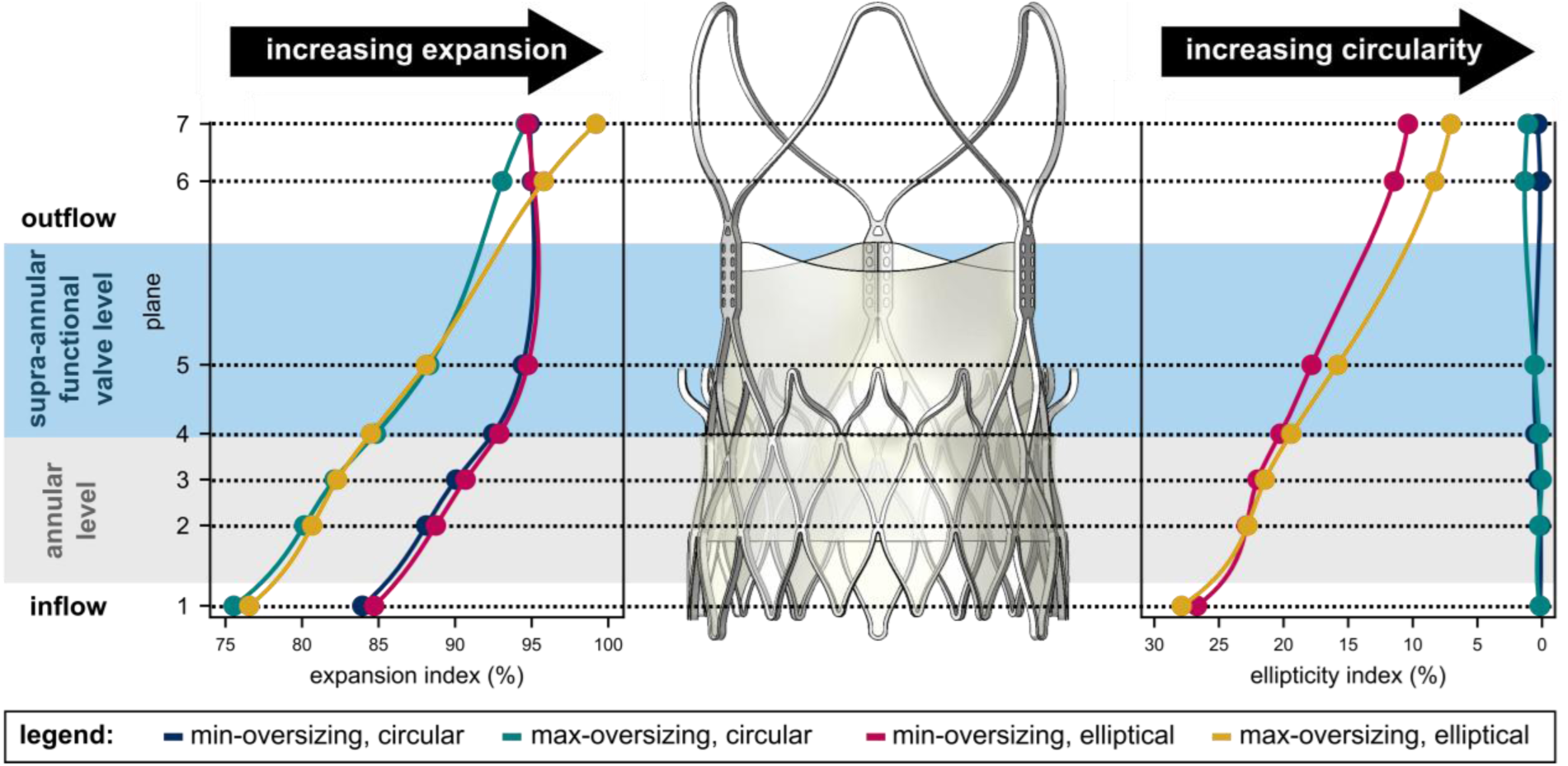
Stent frame expansion and ellipticity following supra-annular deployment calculated using measured device diameter at plane, showing ‘stent-frame decoupling’. The inflow (plane 1) exhibits the reduced expansion and increased ellipticity. Expansion and circularity gradually increases across the annular level (plane 1-4) to the supra-annular functional valve level (plane 4-5). The outflow (plane 6-7) maintains a more expanded, circular configuration.

#### 2.2.1 Leaflet mechanics

To analyze mechanical stress throughout the leaflet, we divided each leaflet into distinct subsections (Figure 6(a)); (1) commissures, (2) upper belly, (3) central belly (4) lower belly and (5) attachment edge. We then quantify the following: (1) von Mises stress, (2) maximum principal strain, (3) coaptation area and (4) pinwheeling index, comparing differences across cases of THV expansion and ellipticity.

#### 2.2.2 Hemodynamic wall shear stress indices

We analyze flow at the leaflet-blood boundary through quantification of hemodynamic wall shear stress (WSS) metrics, which may be linked to the pathology of SVD [12, 15, 16]. This includes the calculation of time-averaged wall shear stress (TAWSS), oscillatory shear index (OSI) and relative residence time (RRT) using the equations below respectively:

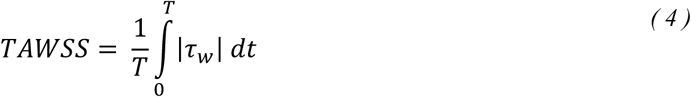

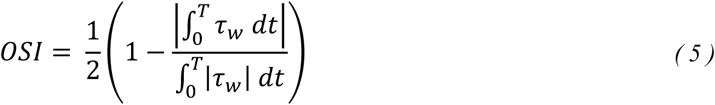

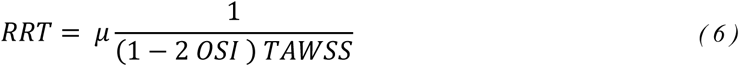

where *τ*_*w*_ is the WSS and *μ* is the dynamic viscosity. These hemodynamic WSS indices are compared across the leaflet regions using the same subsections as defined in Section 2.2.1.

#### 2.2.3 Hemodynamics downstream of the THV

To examine the impact of expansion and ellipticity on flow downstream of the THV, we compute the instantaneous viscous shear stress (VSS), which is linked to platelet damage and hemolysis [19, 38], using equation 4 below:

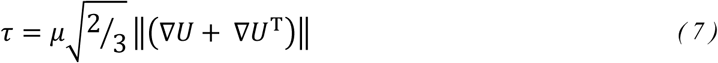

where *τ* is the VSS in Pascal (Pa), ∇*U* is the velocity gradient and *μ* is the dynamic viscosity (N.s/m^2^). We then create a probability density function of this variable, which was displayed in a semi-log scale.

#### 2.2.4 Stent fatigue

We also examined the impact of device deployment on stent fatigue, by reporting stent deflection across each deployment configuration [22, 39, 40]. We extracted these parameters from the second cardiac cycle, and performed a stent fatigue analysis, as per criteria proposed by both Pelton et al. [39] and Cao et al. [40], which describes the fatigue resistance of nitinol in relation to alternating and mean strains based on fatigue testing conducted 10 million and 400 million cycles respectively. This involves obtaining the alternating strain (*ɛ*_*a*_) at each element in the stent and plotting this against (1) the position of the element along the height of the stent and (2) the mean strain (*ɛ*_*m*_), using equation 5 and 6 below;

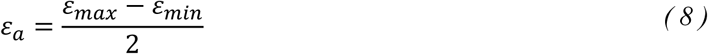

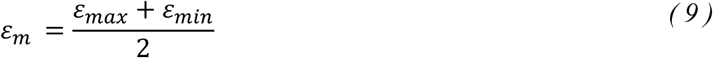

The minimum fatigue factor of safety (FOS) combines the Goodman criteria from Pelton et al. and Cao et al. [39, 40]. This combined limit, as shown in Figure 12(a), takes the minimum alternating strain limit along the mean strain from each study, to create a more stringent, combined fatigue limit to describe the fatigue resistance of nitinol relative to previous experimental testing [39, 40].

## 3 Results

### 3.1 Supra-annular THV deployment results in stent-frame decoupling

To investigate the occurrence of ‘stent-frame decoupling’ in supra-annular self-expanding THVs [34], we characterized device expansion and ellipticity at specific planes along the length of stent frame (inflow, annular, supra-annular functional valve, outflow), see Figure 4. THV expansion was calculated based on the ratio of the predicted device size at the planar level (after the computational simulation of deployment) to the measured nominal device size (not the labelled device diameter). Across all deployment configurations (circular/elliptical, min-oversizing/max-oversizing), frame expansion was the lowest at the inflow (plane 1) and increased from the inflow to the supra-annular region, where the functional valve region is located (Figure 4). In cases of maximal oversizing, where the perimeter of the annulus was the smallest recommended for the XL-sized device, stent frame expansion was less than 90% between plane 1 and 5. High frame ellipticity (∼27%) was found at planar level 1 for both the ‘min-oversizing, elliptical’ and ‘max-oversizing, elliptical’ configurations. This was associated with a gradual decrease in ellipticity at the functional valve region (∼20% - plane 4/5) and outflow (∼9% - plane 6/7). Of note, the idealized elliptical annulus models were designed to represent worst-case ellipticity across the supra-annular valvular level. Thus, we observe stent-frame decoupling of the self-expanding THV, with frame expansion and circularity increasing from the frame inflow to the supra-annular functional valve region. This has been noted in clinically studies of supra-annular, self-expanding devices [33, 34], thereby validating the clinical-relevance of the deployment configurations investigated as part of this study.

### 3.2 Maximum THV oversizing reduced tissue stresses near the leaflet commissures, but altered hemodynamics causing early flow separation and increased shear stresses downstream

Contour plots of the maximum principal strain in the THV tissue leaflets under peak diastolic loading (t = 0.430 s) for each deployment configuration are shown in Figure 5(a). We examined the stress distribution across five distinct regions of the leaflet (commissures, upper belly, central belly, lower belly and attachment edge) across the entire cardiac cycle, as shown in Figure 6(a). Across all deployment configurations, relatively high von Mises stresses were observed in the lower belly and attachment edge of the tissue leaflets during systole (Figure 6(b), Supplementary Materials Figure S1). However, the von Mises stress was consistently higher throughout the leaflet during diastole, when compared to systole, with peak stress at (t = 0.430 s) occurring at the commissure region of the leaflet for all deployment cases. At peak diastole, shown in Figure 6(c), maximum-oversizing (‘circular’ and ‘elliptical’) reduced stresses in the commissure region of the leaflet by 4.9% (to 1.06 MPa) and 8.4% (to 1.05 MPa) when compared to minimum-oversizing counterparts. Across all deployment configurations, higher stresses were observed in the lower belly region at peak diastole when compared to the upper belly, central belly and attachment edge (Figure 6(c)). However, in these regions, maximum-oversizing resulted in a stress redistribution, where von Mises stress magnitudes decreased in the upper and central belly region and increased in the lower belly and attachment edge, relative to the minimum-oversizing case.

**Figure 5:**
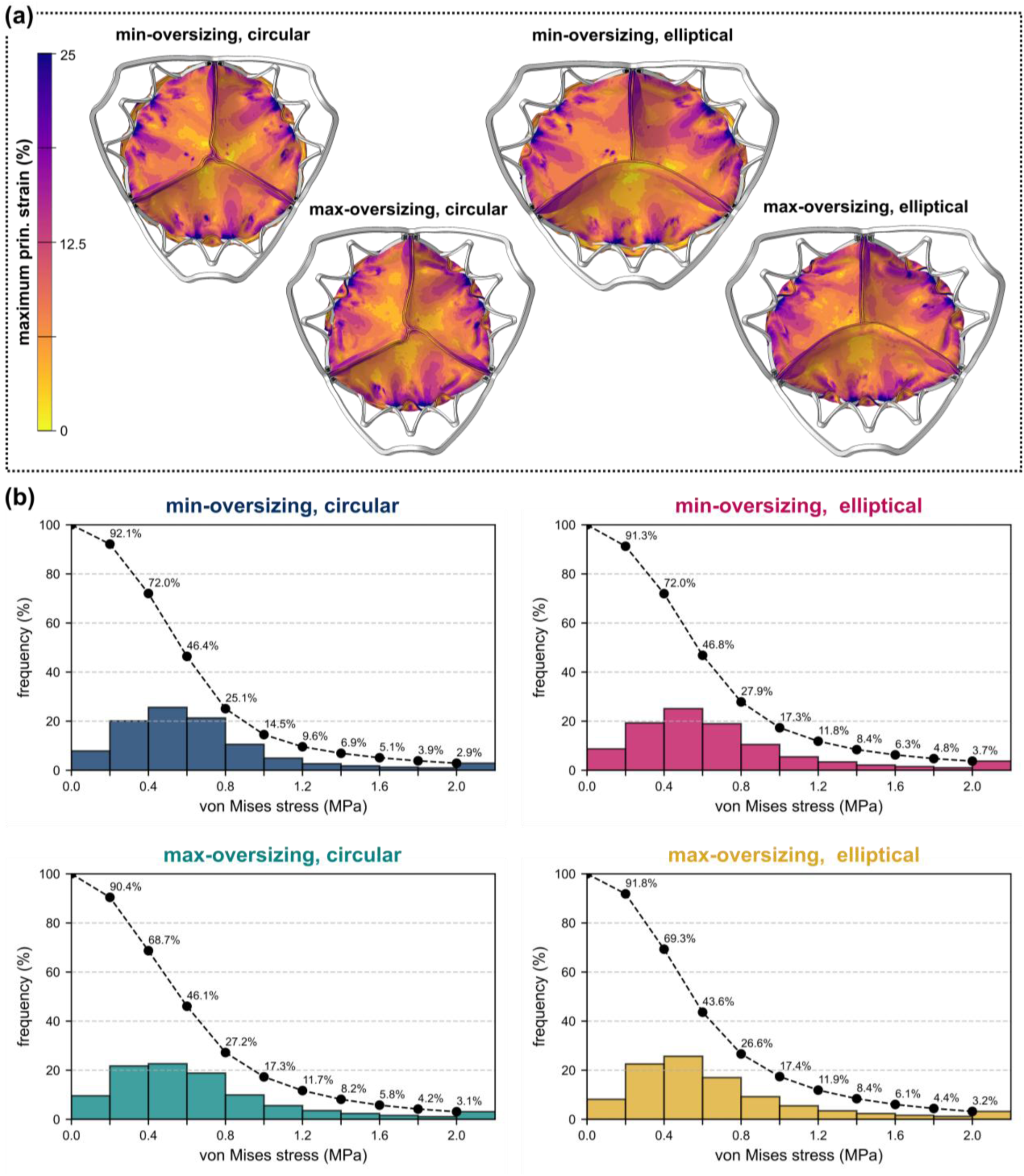
Stress-strain distribution through leaflets of the THV. (a) Contour plots of the maximum principal strain at peak diastolic loading (t = 0.430 s). (b) Distribution of the von Mises stress throughout leaflet as a function of percentage volume, also showing the reverse cumulative von Mises stress (black line), for each deployment configuration.

**Figure 6:**
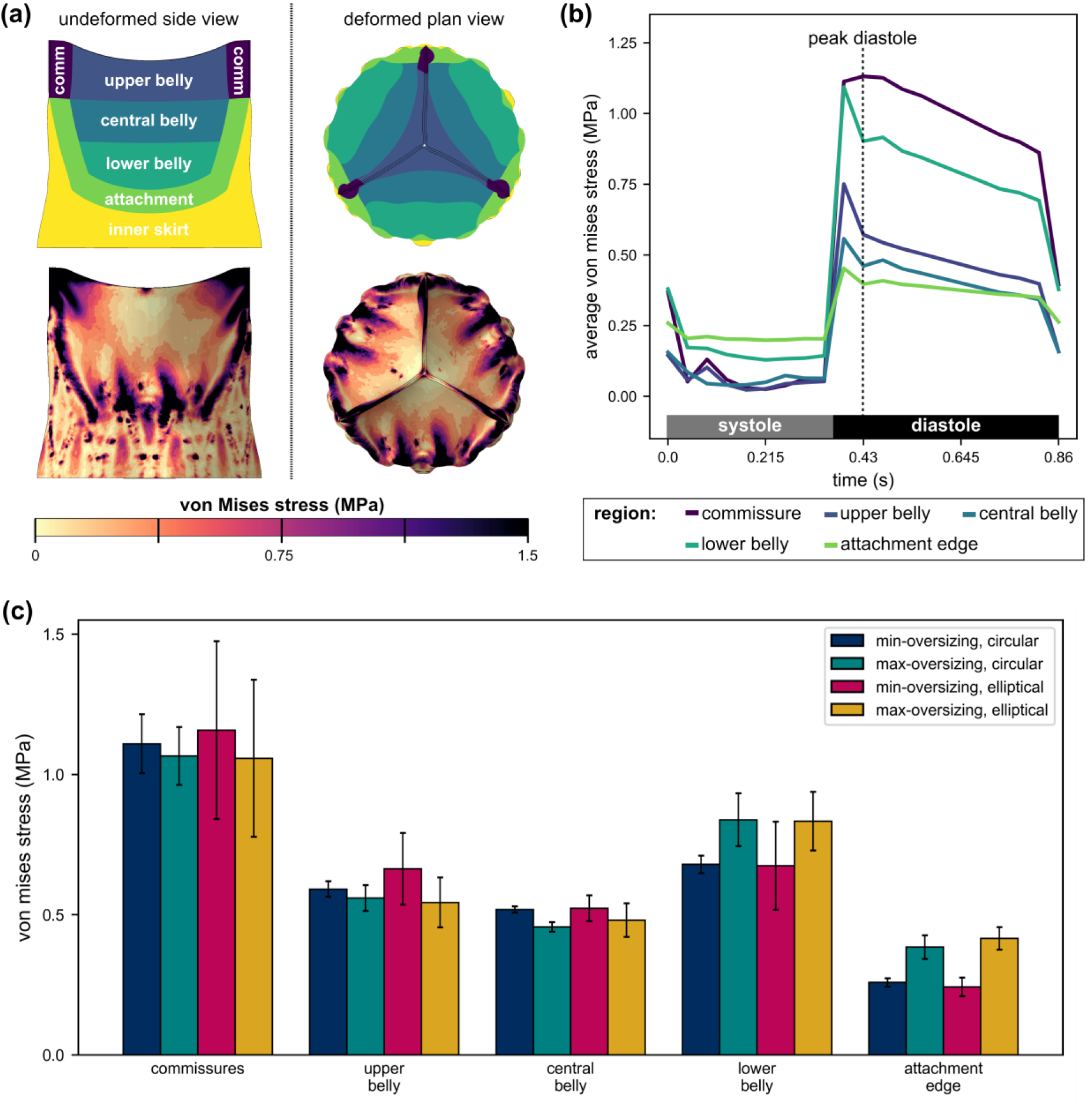
Sub-regional leaflet stress distribution. (a) Schematic of undeformed (-left) and deformed (at peak diastole – right) leaflet sub-regions with for calculation of average von Mises stress in each region throughout the cardiac cycle (b) Average von Mises stress in the defined leaflet sub-regions across the entire cardaic cycle for ‘max-oversizing, circular’ case (other cases shown in Supplementary Figure S1). (c) Histogram comparing the average and standard deviation of von Mises stress across each leaflet at peak diastole (t = 0.430 s) for all deployment configurations.

Oversizing of the THV altered device dynamics by increasing average coaptation area, see Figure 7(a). The average coaptation area of the leaflets at peak diastole (Figure 7(b)) was 19.3% and 18.0% in the ‘min-oversizing, circular’ and ‘min-oversizing, elliptical’ deployment configurations, when compared to 23.9% and 24.3% in the ‘max-oversizing, circular’ and ‘max-oversizing, elliptical’ cases respectively. The THV exhibited clear pinwheeling across all deployment configurations, as evident in Figure 7(a). In the ‘min-oversizing, circular’ and ‘min-oversizing, elliptical’ annuli, the PI was equal to 2.7% and 3.1% respectively, which increased to 4.8% and 4.6% in the ‘max-oversizing, circular’ and ‘max-oversizing, elliptical’ annuli respectively. Thus, our model predicts that pinwheeling increases with oversizing and reduced THV expansion, which aligns with previous studies [29] and *in vitro* testing results described in [24]. We quantified the impact of pinwheeling on leaflet maximum principal strain, as an indicator of long-term device durability, at the tip of the leaflet, as shown in Figure 7(d). We predicted that the mean maximum principal strain at the leaflet tip increased in the ‘max-oversizing, circular’ deployment configuration (6.1%), when compared to the ‘min-oversizing, circular’ annulus (4.7%).

**Figure 7:**
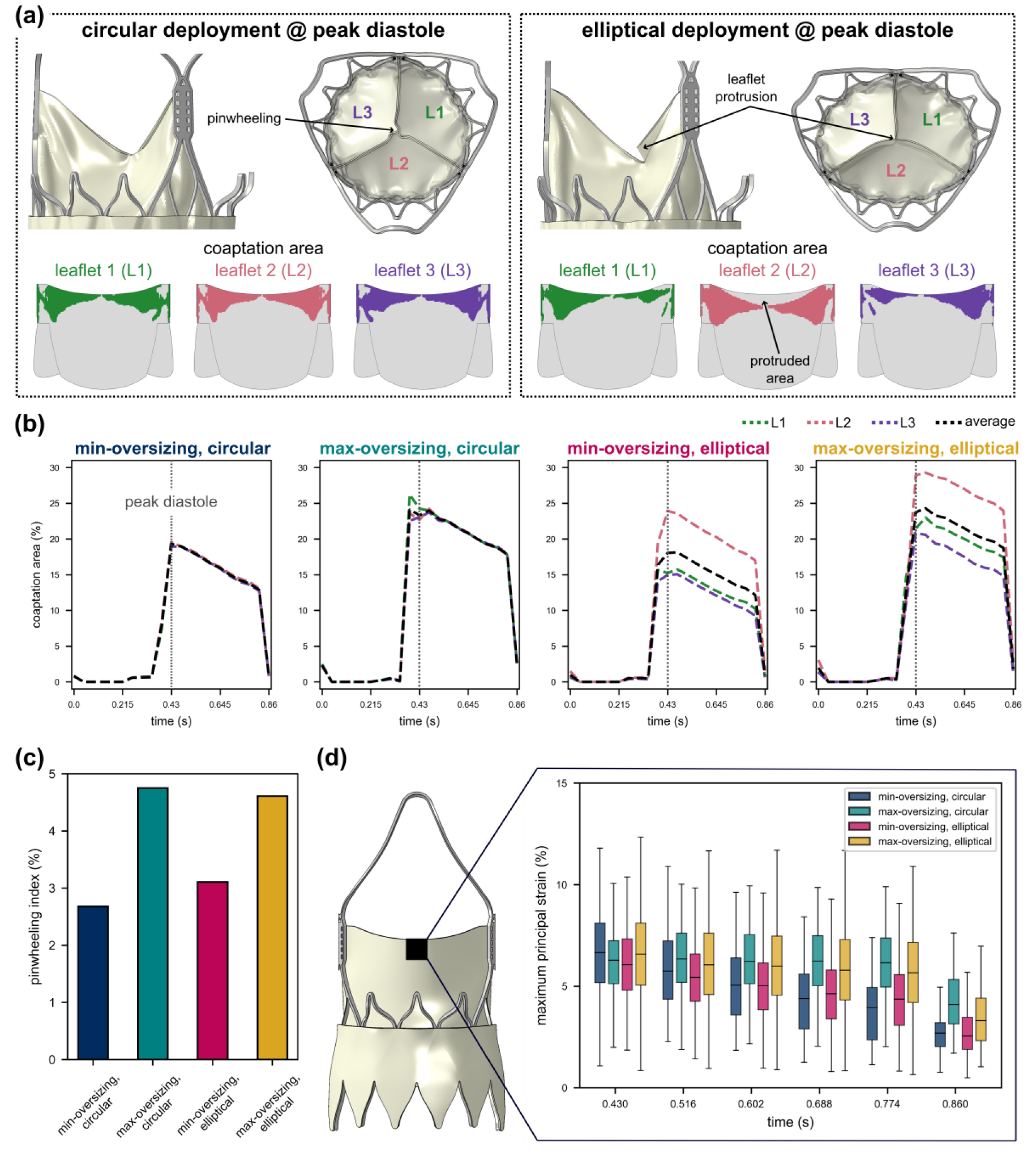
Leaflet coaptation indices. (a) Coaptation and pinwheeling of THV deployed in circular (left) and elliptical (right) annulus, showing coaptation area (coloured by leaflet) and leaflet protrusion evident in elliptical deployment configurations. (b) Quantification of coaptation area during the cardiac cycle showing average, leaflet 1 (L1 - green), leaflet 2 (L2 – pink) and leaflet 3 (L3 – purple) coaptation area. (c) Pinwheeling index at peak diastole (t = 0.430 s) across each deployment configuration. (d) Maximum principal strain at the leaflet tip (black region) during coaptation.

Table 3 shows hemodynamics WSS indices assessed as part of this study across all deployment configurations. We found, as expected, that the surface-averaged TAWSS was greater on the ventricular surface of the leaflets, when compared to the aortic surface across all deployment configurations (Figure 8(b)). Our predicted surface-averaged TAWSS values across the aortic (0.4 – 0.7 Pa) and ventricular (2.4 – 3.2 Pa) surfaces respectively, which were in keeping with previous studies [12, 41]. Oversizing increased the TAWSS on the aortic surface (Figure 8(b)), but decreased TAWSS on the ventricular surface, with a noticeable reduction in the upper and central belly regions (Supplementary Figure S2). This was caused by early mainstream flow separation (see ‘max-oversizing, circular’, ‘max-oversizing, elliptical’ in Figure 8(a)), when the boundary layer detached from the leaflet surface, which resulted in recirculation, vortical flow and increased OSI on the ventricular surface (Figure 9(a)). On the aortic surface, the surface-averaged OSI was reduced with device oversizing, particularly in the central belly, lower belly and attachment edge regions (Supplementary Figures S3). In the ‘max-oversizing, circular’ and ‘max-oversizing, elliptical’ configurations, the median RRT across the aortic surface was equal to 0.0195 s and 0.0147 s respectively, which was lower than that for fully-expanded configurations, see Table 3.

**Figure 8:**
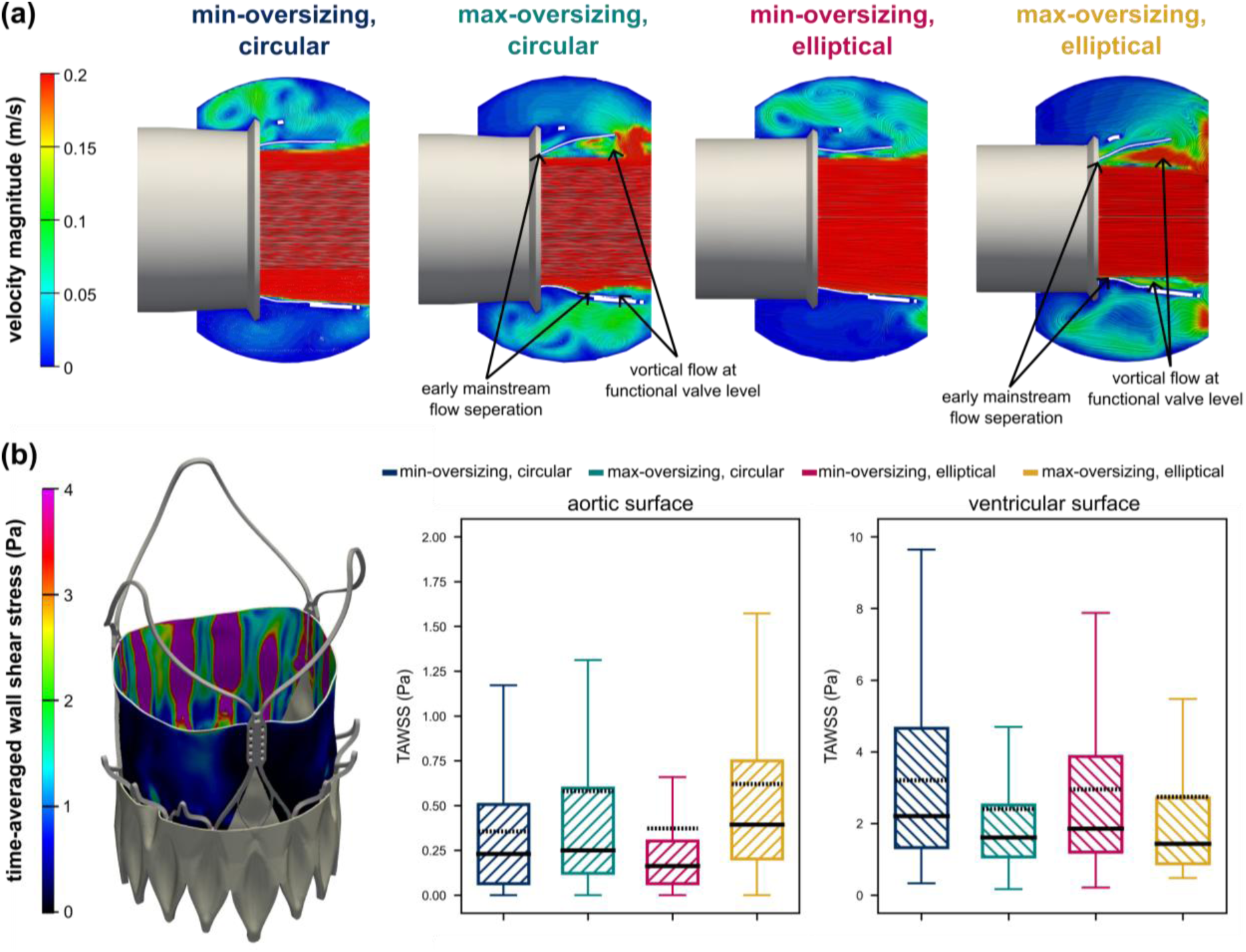
Maximum-oversizing alters hemodynamics near the THV leaflet; (a) Velocity contours at peak systole showing hemodynamics around the THV, with early main stream flow separation and vortices at functional valve level with device oversizing and reduced expansion. (b) Time-averaged wall shear stress (TAWSS) on the aortic and ventricular surface of the leaflets showing median and mean (dashed line).

**Figure 9:**
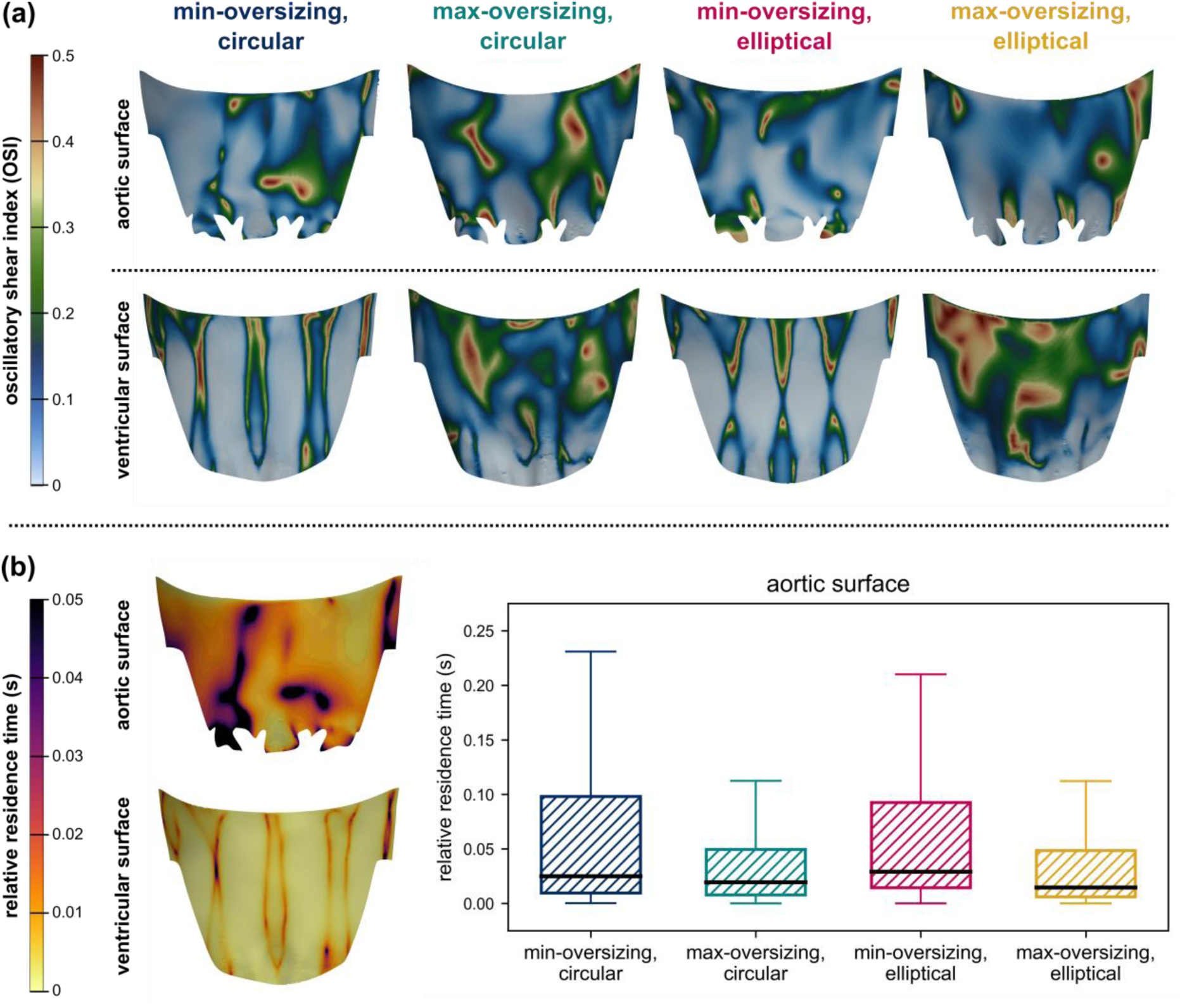
THV oversizing and ellipticity altered hemodynamic WSS indices; (a) increased oscillatory shear index (OSI) on the ventricular surface in cases of maximum-oversizing and (b) reduced relative resident time (RRT) on the aortic surface.

**Table 3:** Summary of surface-averaged hemodynamic indices of structural valve degeneration.

| Hemodynamic index of SVD | Surface | Min-oversizing, circular | Max-oversizing, circular | Min-oversizing, elliptical | Max-oversizing, elliptical |
| --- | --- | --- | --- | --- | --- |
| TAWSS | aortic | 0.356 | 0.582 | 0.372 | 0.621 |
|  | ventricular | 3.206 | 2.408 | 2.956 | 2.747 |
| OSI | aortic | 0.131 | 0.118 | 0.107 | 0.107 |
|  | ventricular | 0.085 | 0.149 | 0.086 | 0.114 |
| RRT | aortic | 0.025 | 0.020 | 0.029 | 0.015 |
|  | ventricular | 0.002 | 0.004 | 0.002 | 0.004 |

The instantaneous VSS downstream of the THV (outflow section of the pulse duplicator) is shown in Figure 10(b) for all deployment configurations. We predicted that oversizing increased the likelihood of increased instantaneous VSS, when compared to minimum-oversizing (also evident in Figure 10(a)). The range of VSS reaches a maximum (*σ*_99_) of 30.0 Pa and 36.4 Pa in the ‘min-oversizing, circular’ and ‘max-oversizing, circular’ deployment configurations respectively (Table 4). Furthermore, in the ‘max-oversizing, circular’ case, a greater portion of the flow field is exposed to higher VSSs (> 10 Pa) which may lead to platelet damage [19, 38], when compared to the ‘min-oversizing, circular’ case (1.83% vs 1.52%) respectively.

**Figure 10:**
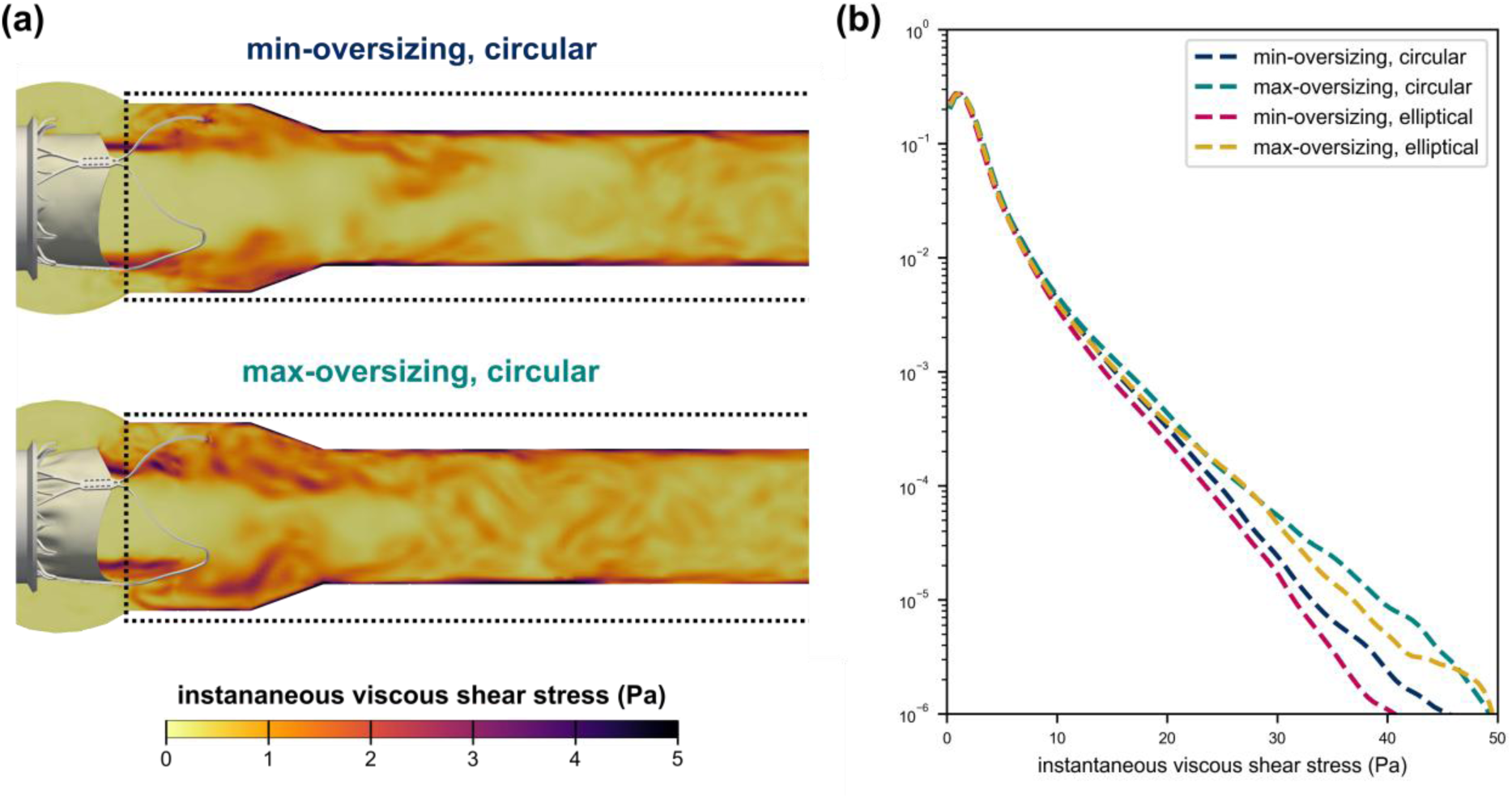
Instantaneous viscous shear stress (VSS) and implications for thrombogenicity. (a) Contours of the instantaneous VSS for the ‘min-oversizing, circular’ and ‘max-oversizing, circular’ case showing outflow section (black hashed line) used for probability density function calculation. (b) Probability density function (PDF) of instantaneous VSS distribution in the flow downstream of the THV throughout a complete cardiac cycle in semi-log scale.

**Table 4:**
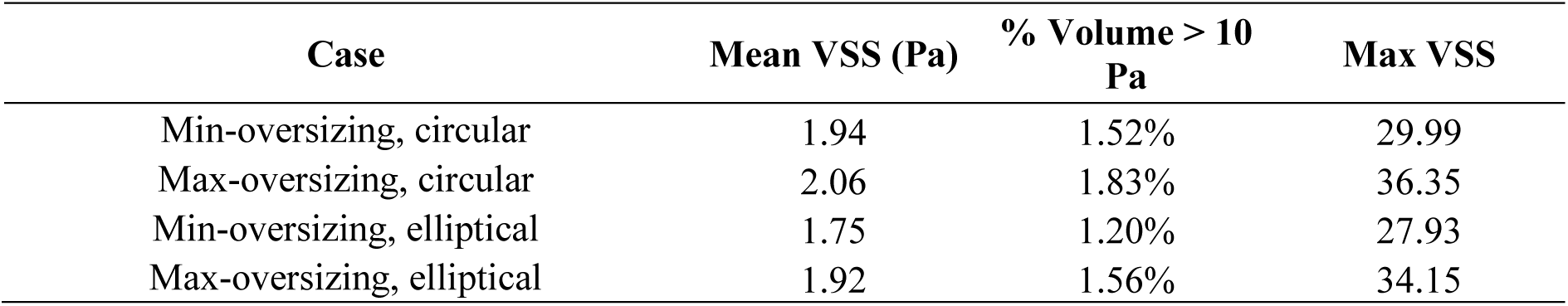
Instantaneous VSS downstream of the THV.

Oversizing reduced average stent deflections (1.21 mm) by over 35% in the ‘max-oversizing, circular’ deployment configuration, when compared to the ‘min-oversizing, circular’ case (1.95 mm). Given that such cyclical stent deflections may present a risk of fatigue-related failure, we also report the alternating and mean strains along the stent. Across all deployment configurations, the maximum alternating strain occurred at the commissure region of the stent (x = 0 mm - Figure 11(a)). However, we found that alternating strains decreased throughout the stent when the device was oversized, particularly at points where the leaflet is sutured to the stent at the upper crown (-18 < x < -14 mm) and free cells (y = -27 mm). Across all deployment configurations, the maximum mean strain occurred at the upper crown of the stent (-18 < x < -12 mm - Figure 11(c)). Oversizing increased mean strains throughout the stent, particularly in regions where the stent is constrained at the annular level (-15 < x < -35 mm), where the maximum mean strain was equal to 3.79% for both the ‘max-oversizing, circular’ and ‘max-oversizing, elliptical’ cases respectively. Thus, the minimum fatigue FOS in the ‘max-oversizing, circular’ deployment configuration occurred at the annular level (x = -20 mm – Figure 12(c)). In the ‘min-oversizing, circular’ deployment configuration, the maximum deflecting commissure post P1 was most at risk of fatigue failure, due to the large alternating strains exhibited by the THV stent at this point during cyclical loading of the leaflet (Figure 12(b)).

**Figure 11:**
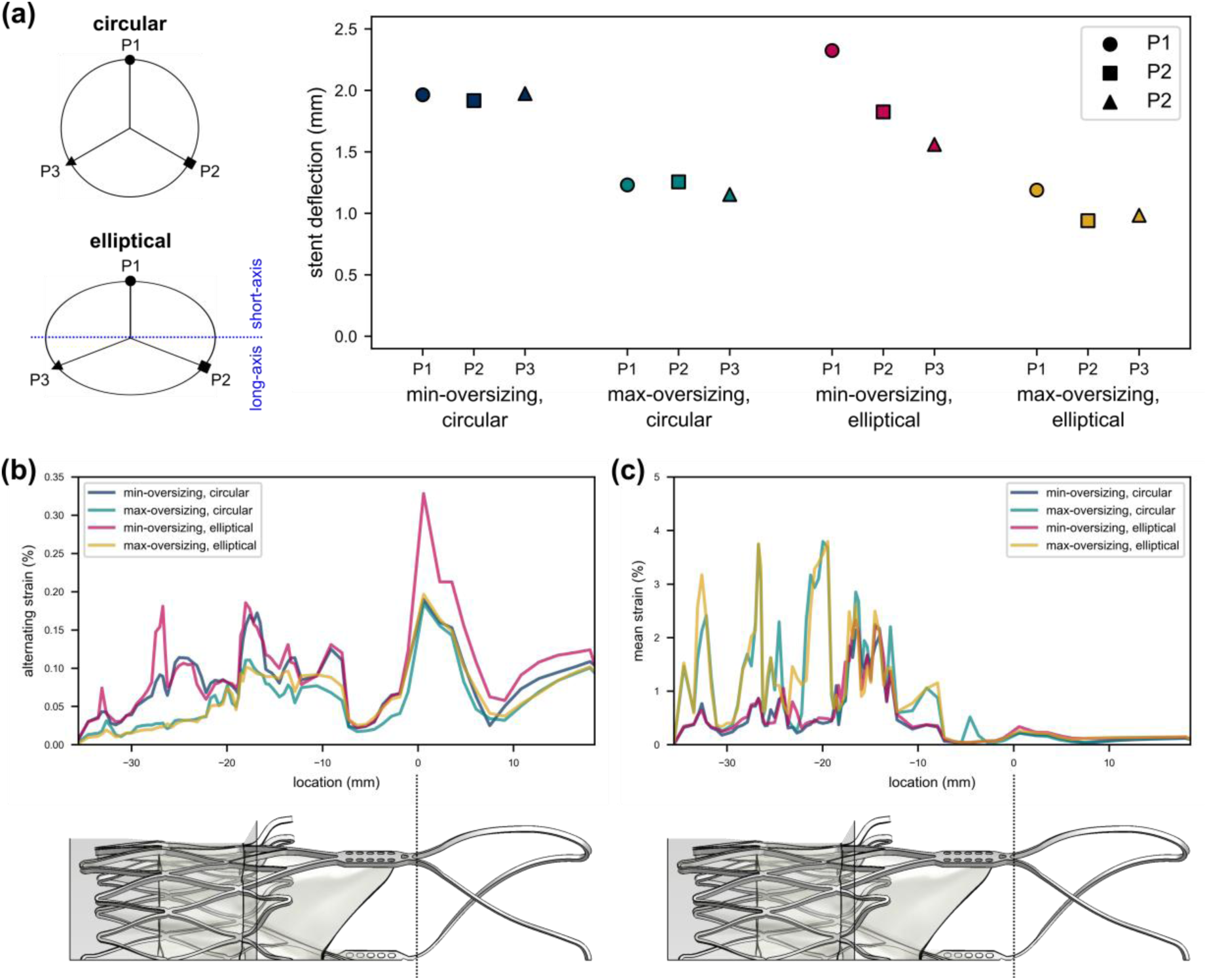
Synergistic impact of THV deployment and stent deformability during cardiac cycle on stent deflections and alternating/mean strains. (a) Stent deflection, a measure of the radial deflection of each stent commissure between systole and diastole. (b) Maximum alternating strains extracted from all elements along the length of the stent. (c) Maximum mean strains along extracted from all elements along the length of the stent.

**Figure 12:**
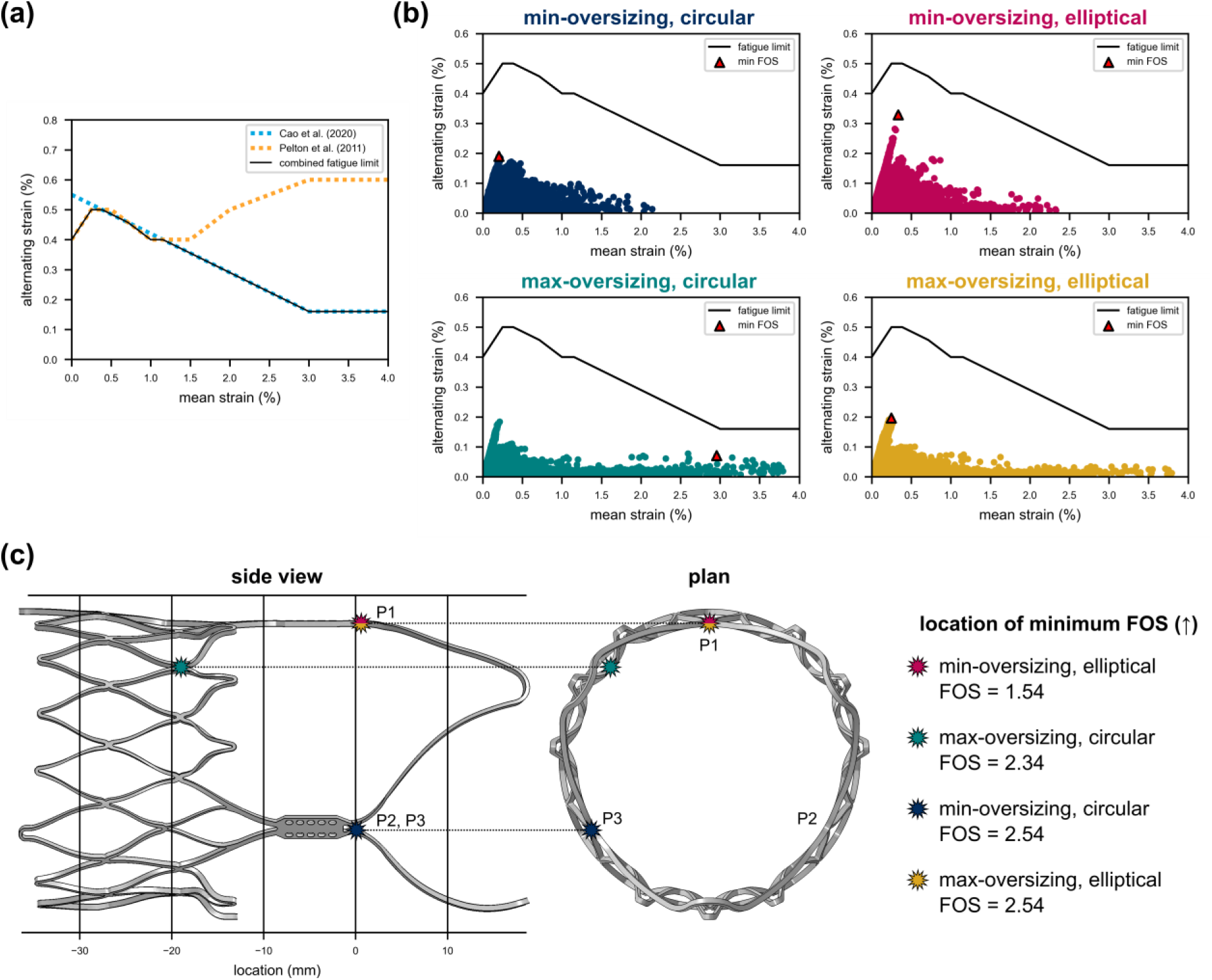
Stent fatigue predictions. (a) Combined fatigue limit using Goodman criteria from Cao et al. (2020) and Pelton et al. (2008). (b) Constant-life fatigue diagrams across each of the deployment conditions showing element with minimum fatigue factor of safety (FOS). (c) Location of minimum fatigue FOS for each deployment configuration.

### 3.3 Elliptical deployment induced heterogenous THV stent deflections, coaptation mismatch and variable leaflet stress distributions

Elliptical deployment did not notably increase the average von Mises stress in the tissue leaflet subregions at peak diastole (Figure 6(c)). The maximum von Mises stress occurred at the commissure region, where average values across all three leaflets were equal to 1.16 MPa and 1.06 MPa for ‘min-oversizing, elliptical’ and ‘max-oversizing, elliptical’ cases, respectively. However, elliptical deployment resulted in heterogenous leaflet stress distributions, where stress was increased in one leaflet subregion compared to the same subregion of another leaflet, evident by the increased standard deviation in Figure 6(c). In the ‘min-oversizing, elliptical’ case, the mean von Mises stress in the commissure region was equal to 1.33 MPa, 0.836 MPa and 1.305 MPa across leaflet 1 (L1), leaflet 2 (L2) and leaflet 3 (L3) respectively (Table S1 in Supplementary Materials X), when compared to 1.02 MPa, 1.094 MPa and 1.22 MPa in the circular case. A similar pattern was observed in the upper belly region, where the average von Mises stress of L1, L2 and L3 was equal to 0.70 MPa, 0.60 MPa and 0.69 MPa respectively, which were both higher and more variable than in the circular case. However, THV ellipticity did not alter the volume of leaflet tissue experiencing higher stresses, which was similar across all deployment configurations (Figure 5(b)).

Our model predicted that THV ellipticity is associated with coaptation mismatch. For both the ‘min-oversizing, elliptical’ and ‘max-oversizing, elliptical’ configurations, L2, located parallel to the long-axis of ellipticity, exhibited a larger coaptation area than L1 and L3, see Figure 7(a). L2 also protruded above other leaflets during diastole (grey area above coaptation region in L2), which has been reported in previous *in silico* and *in vitro* studies [7, 42]. Although, this did not alter the maximum principal strain at the leaflet tip (Figure 7(c)), it may have caused the relative increase in von Mises stress noted in the upper belly region of L2 and L3, when compared to L1, at peak diastole.

Elliptical deployment did not impact the TAWSS, OSI and RRT, when compared to circular deployment, but reduced the instantaneous VSS downstream of the THV, see Table 4 and Figure 10(b).

We predicted heterogeneous stent deflection across cases of elliptical THVs, whereas in circular cases deflection was similar across commissure posts (Figure 11(a)). In the ‘min-oversizing, elliptical’ and ‘max-oversizing, elliptical’ annuli, maximum stent deflection occurred at commissure post P1, which was positioned along the short-axis of ellipticity. The deflection of commissure stent post P1 in the ‘max-oversizing, elliptical’ deployment configuration was half that of the ‘min-oversizing, elliptical’ case. Furthermore, P2 and P3 (along the long-axis) exhibited deflections that were approximately 80% of P1 deflection for both ‘min-oversizing, elliptical’ and ‘max-oversizing, elliptical’ deployment. This commissure post had the lowest fatigue FOS for both min- and max-oversizing elliptical deployment configurations, equal to 1.54 and 2.54 respectively (Figure 12(b)).

## 4 Discussion

In this study, we investigated the impact of THV oversizing and ellipticity on biomechanical and hemodynamic indices of structural degeneration and thrombogenicity, using a validated *in silico* framework [24]. This is the first computational study to examine the synergistic impact of stent deformation, due to pulsatile loading of the bioprosthetic leaflets during cardiac function, and THV deployment configuration. We found that supra-annular deployment of a self-expanding THV resulted in ‘stent-frame decoupling’, which was associated with a gradual increase in frame expansion and circularity from the inflow to the outflow. Elliptical deployment of a self-expanding THV induced heterogenous stent deformations during pulsatile loading, which had altered device closure kinematics, resulting in coaptation mismatch and heterogeneous leaflet stress distributions. However, peak leaflet stresses observed were notably less than previous studies, which did not account for either stent deformability or ‘stent-frame decoupling’. Stent posts along the short-axis of ellipticity experienced the greatest level of deflection during pulsatile loading, which may increase the risk of stent fatigue failure at these points. Our models also showed that maximum-oversizing reduced expansion at the frame inflow and functional valve region (< 90%), decreased stent deflection and leaflet stresses at the commissures, but increased stress magnitudes within the lower belly and attachment edge leaflet regions. Although ellipticity did not have a notable impact on device hemodynamics, oversizing reduced the TAWSS and increased the OSI on the ventricular surface of the leaflet, due to early flow separation, and increased VSS downstream of the THV. The findings of this study provide insight into the impact of device oversizing and ellipticity on the long-term function and durability of supra-annular, self-expanding THVs.

There are a number of limitations of this study that require consideration. Firstly, we did not account for coupled multi-physics (solid, fluid) interactions between the device and adjacent fluid flow. However, previous comparative studies utilizing discrete FE/CFD and FSI simulation methods have reported similar von Mises stress and WSS distributions [12, 16, 41, 43]. Having said this, systolic leaflet fluttering, an identifier of early morphological SVD [8], may only be captured using FSI approaches. Ongoing work aims to address this limitation by developing a fully-coupled FSI model of the self-expanding THV. Secondly, the strains experienced by the leaflets during the cardiac cycle exceeded the strain range to which glutaraldehyde-fixed porcine pericardium tissue was tested previously [35]. Thirdly, our stent fatigue FOS is based on combining constant-life diagrams of nitinol, which cannot predict the time to failure. However, this approach may be used to compare alternating and mean strains across multiple cases of THV stent deformation based on our current knowledge of nitinol fatigue. Next, we assumed flow was laminar, whereas intraventricular flow undergoes periodic transformations to laminar-transitional in certain phases of the cardiac cycle. Moreover, our models excluded coronary flow, which may increase wash-out, reduce flow stasis and alter WSS patterns [30, 44]. Finally, we used idealized annulus models to examine THV deployment, whereas the native aortic valve is typically heavily calcified which may cause further device underexpansion, ellipticity and asymmetry. Future studies will conduct further model applicability analyses to replicate the TAVI procedure in virtual, patient-specific cohorts, which may be applied to study retrospective cases of structural degeneration and thrombogenicity.

THVs are prone to early failure due to SVD, which may be caused by the interaction between the host environment and biomechanics. In particularly, it is postulated that regions of high mechanical stress are subject to calcification. In this study, we showed that high stress occurs at the leaflet commissures at peak diastole, which aligns with previous studies and failure regions observed *in vivo* [7, 11, 26]. Although average stresses were similar, THV ellipticity resulted in increased stresses (+18.2%) at the commissure of the leaflet located along the short-axis of ellipticity when compared to the same region during circular deployment. Although much higher stress increases (+117% , +2161%) have been reported at leaflet commissures of eccentrically deployed THV’s [7, 26], previous studies did not account for stent deflection during pulsatile loading, and assume stent rigidity, which increases peak leaflet stress magnitudes [45]. Here, we incorporated both stent deformability and observed ‘stent-frame decoupling’ – a phenomenon observed clinically [33, 34] but not previously explored computationally. We report that the THV stent exhibited asymmetric deflections following elliptical deployment, which induced heterogenous leaflet stress distributions and coaptation mismatch, where one leaflet protruded above the others, which has also been noted in studies of SAVR function [42]. Despite this, we found that THV ellipticity did not notably increase the volume of leaflet tissue experiencing higher stresses, which contrasts with previous studies [7, 26] that assumed stent rigidity during cardiac function, or investigated intra-annular deployment and uniform stent expansion. Despite being a prominent issue, particularly for self-expanding TAVI, the impact of THV ellipticity and implications for long-term valve function have not been thoroughly examined in clinical studies. Although THV ellipticity has been associated with hypo-attenuated leaflet thickening (HALT) at one-year (a predictor of SVD [46]) [47, 48], another study reported no significant differences in device ellipticity between patients with and without SVD at 5-year follow-up [49]. In this study, we showed that idealized elliptical deployment does not notably impact hemodynamics around or downstream of the THV, which is in keeping with a previous study [50]. Furthermore, this study suggests that both (1) heterogeneous stent deflections noted in cases of THV ellipticity and (2) ‘stent-frame decoupling’ may reduce the impact of elliptical deployment on leaflet stress magnitudes, thereby potentially improving the long-term durability of these devices.

In this study, we found that maximum-oversizing of the THV in an idealized, bench-top model resulted in incomplete device expansion (< 90%) from the inflow to the supra-annular functional valve level. We further found that maximum-oversizing altered leaflet dynamics, resulting in a stress redistribution – decreasing stresses in the commissure, upper belly and central belly regions, and increasing stresses in lower belly and attachment edge regions. Additionally, we report that maximum-oversizing increased leaflet coaptation area and pinwheeling, which has long been linked to reduced device durability [25, 29]. Indeed, we found that pinwheeling was directly correlated with increased maximum principal strain at the leaflet tip, which contrasts from recent findings that report no relationship between pinwheeling and leaflet stress [45]. Although it is important to note that the previous study [45] utilized shell-based elements, which may not be adequate for capturing leaflet transmural behavior [51]. Furthermore, recent *in vitro* studies report tearing at the leaflet tip of a SAVR device [42], which further supports our findings. We also observed altered hemodynamics around and downstream of the THV, associated with varying degrees of device expansion resulting from different levels of intentional oversizing. On the aortic surface of the leaflets, maximum-oversizing increased the TAWSS and decreased the RRT. Interestingly *in vitro* studies of supra-annular THVs showed enhanced neo-sinus washout in similar idealized cases of incomplete device expansion (< 90%) [52], which would be expected with decreased residence time and may reduce stagnation induced thrombosis on the aortic surface. However, thrombi have been observed on both leaflet surfaces of explanted SAVR devices [53, 54], and clot composition may vary due to differences in adjacent surface-specific flow patterns. Our model also predicted that maximum oversizing of THVs is associated with reduced TAWSS and increased OSI on the ventricular surface of the leaflet, which may contribute to long-term hemodynamic valve deterioration [31]. Differences in WSS patterns observed between cases of minimum- and maximum-oversizing were caused by early mainstream flow separation, which resulted in the formation of recirculating vortices within the functional valve region. Recent clinical studies show that greater device oversizing [55] and reduced THV expansion [47] are associated with subclinical leaflet thrombosis and HALT, which are linked to impaired THV durability [56]. Our findings suggest this may be caused by disturbed hemodynamics resulting from maximum device oversizing. Furthermore, maximum-oversizing increased the magnitude of instantaneous VSS downstream, with a larger portion of the flow field exceeding VSS thresholds linked to platelet damage [38]. Aggressive oversizing of self-expanding THVs (beyond the labelled device recommendations) or ineffective pre-dilation of the annulus and/or post dilation may result in device distortion and subsequent underexpansion. Recent studies have shown that underexpansion is associated with a significant increase in stroke and all-cause mortality [57–59], potentially arising from platelet damage caused by elevated VSS. Although underexpansion can be effectively managed through post-TAVI balloon dilatation, it is not commonly identified post-procedure, as its effect on immediate clinical outcomes is not readily apparent [59]. Our results and previous clinical studies highlight the importance of assessing THV expansion, and while post-TAVI CT is the current gold-standard, it is not routinely performed. Recent studies have developed methods to assess THV expansion through fluoroscopy approaches [59], which may be used in real-time to identify patients at risk of increased thrombogenicity and tissue degeneration.

Given the increasing use of TAVI in younger patient cohorts, stent fatigue is also of increasing concern. Here, we present the first study to examine the synergistic effect of THV deployment conditions and pulsatile loading on stent fatigue. We predicted that self-expanding THVs are subject to constant compression from the aortic annulus and cyclical pulsatile loading of the leaflet. In this study, maximum-oversizing reduced average stent deflections, while elliptical deployment caused heterogenous stent deflections. As expected, stent deflection during cardiac loading resulted in large alternating strains at the stent commissure posts in cases of minimum-oversizing, which were at risk of fatigue failure. It is important to note that, although such deformations have been linked to stent fatigue and creep [23], reported strains were well below combined constant-life strain limits, thereby indicating no risk of fatigue failure at 400 million cycles [39, 40]. Furthermore, stent failure has not been reported in medium- and long-term clinical studies of THV function [21, 60]. Although the current study focused on the ACURATE Prime, similar stent deflections have been reported for the CoreValve Evolut PRO (Medtronic, Minneapolis, MN, USA) [23, 32]. Based on findings from this study, we recommend that next-generation THVs should explore the use of flexible stents to reduce the impact of underexpansion and elliptical deployment, whilst also balancing the risk of structural degeneration of the leaflet and stent fatigue.

Recent clinical data suggests that post-deployment geometry may increase thrombogenic risk and contribute to structural degeneration [47, 57–59]. This is the first *in silico* study to consider expansion and ellipticity in the context of clinically observed stent-frame decoupling and stent deformability [23, 24, 32, 34], and we provide an advanced understanding of biomechanical and hemodynamic factors that may lead to structural degeneration and risk of thrombogenic events. Further studies are required to examine the impact of our findings across large clinical cohorts, including (1) explant studies of degenerated THVs, to determine the type and location of structural degeneration, and (2) patient-specific computational approaches, such as those presented in this study, to determine a more advanced outlook on the relationship between biomechanics, thrombogenicity and the onset of structural degeneration.

## 5 Conclusion

This study presents a comprehensive insight into the impact of THV oversizing and ellipticity on device biomechanical and hemodynamic indices of structural degeneration and thrombogenicity. We predicted that elliptical deployment of a THV results in stress heterogeneity throughout the leaflet, which can be reduced by stent deformability and ‘stent-frame decoupling’. Maximum-oversizing of the THV resulted in device expansion less than 90%, which reduced leaflet stresses at commissure and central belly region, increased stress at the lower belly and leaflet tip due to pinwheeling, and altered THV hemodynamics, (flow separation, high viscous shear stress), which may accelerate structural valve degeneration and platelet damage. This study advances our understanding of how THV oversizing and elliptical deployment may influence structural degeneration and thrombogenic risk. This may be used to inform procedural planning, particularly for younger patient-cohorts where increased longevity and degeneration is of increasing concern.

## Data Availability

All data produced in the present study are available upon reasonable request to the authors

## Acknowledgements

The authors would also like to acknowledge the Irish Centre for High-End Computing (ICHEC) for provision of computational facilities and support.

