## Supplementary Materials for "In Silico Modeling of Transcatheter Heart Valve Oversizing and Ellipticity, Part II: Effects on Leaflet Mechanics, Hemodynamics, and Stent Deflection Contributing to Thrombogenic Risk and Structural Degeneration"

### 1 Leaflet von Mises stress distribution

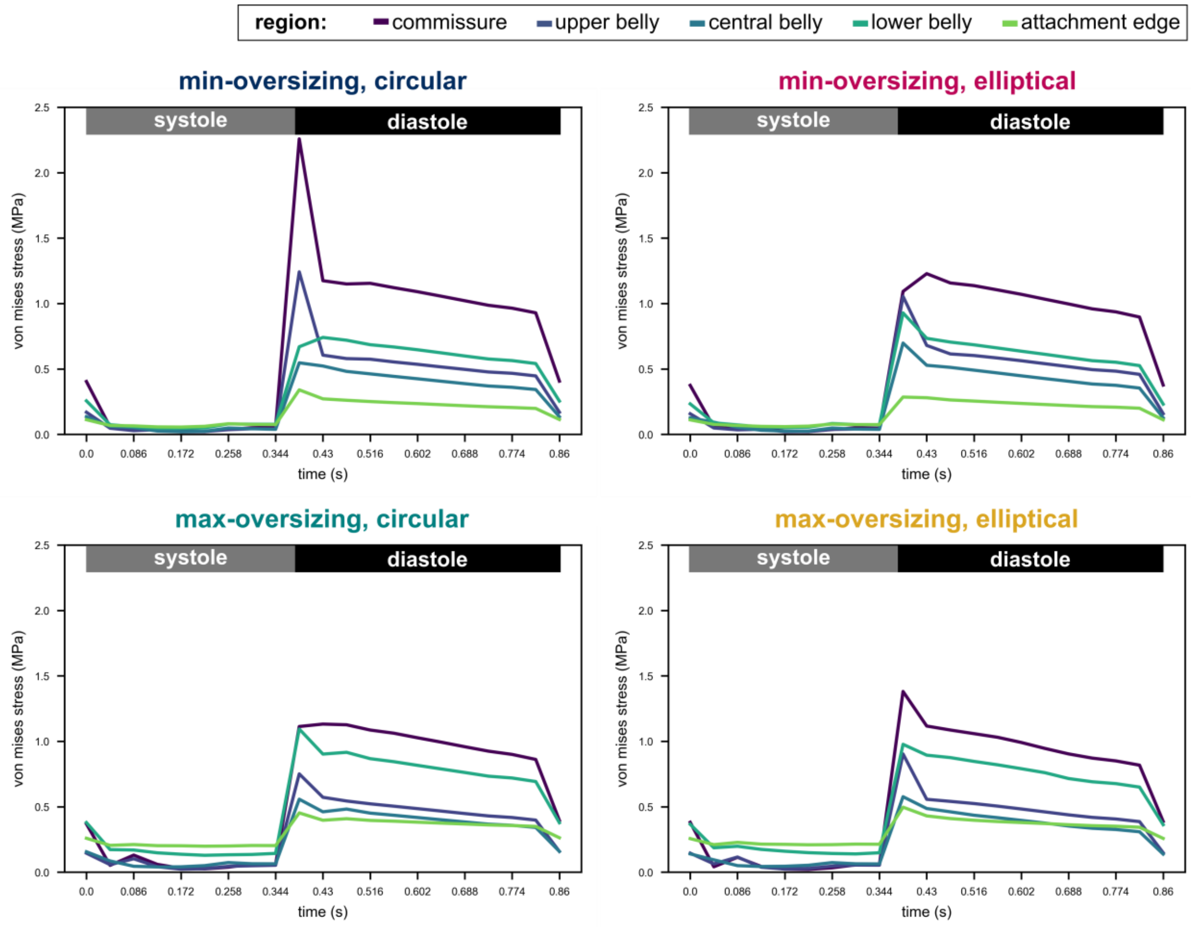


*Figure S1: Average von Mises stress across defined regions of the tissue leaflet for all deployment configurations across the entire cardiac cycle.*

To investigate leaflet-to-leaflet stress variation due to elliptical deployment, we divided each leaflet into two halves, then defined five leaflet subregions (as previously defined in Figure 6(c)) per half leaflet (Figure S2). Elliptical deployment resulted in heterogenous leaflet stress distribution, with peak stresses occurring at the commissure region of the leaflet attached to the stent which exhibited maximum deflection (L1.P1, L3.P1). As shown in Figure S2, commissures parallel to the long-axis (L2), were subject to lower leaflet stresses than leaflets aligned along the short axis (L1,L3). Similar heterogeneous stress distributions and increased peak stress magnitudes with THV ellipticity were noted throughout the belly region of the leaflet. In 'max-oversizing, elliptical' case, the average von Mises stress in the upper belly region of L1, L2 and L3 was 0.70 MPa, 0.60 MPa and 0.69 MPa respectively. These stresses were higher and more varied than those in the upper belly region of the 'max-oversizing, circular' case, which was equal to 0.58 MPa, 0.58 MPa and 0.61 MPa for L1, L2 and L3 respectively (Figure 6(b)).

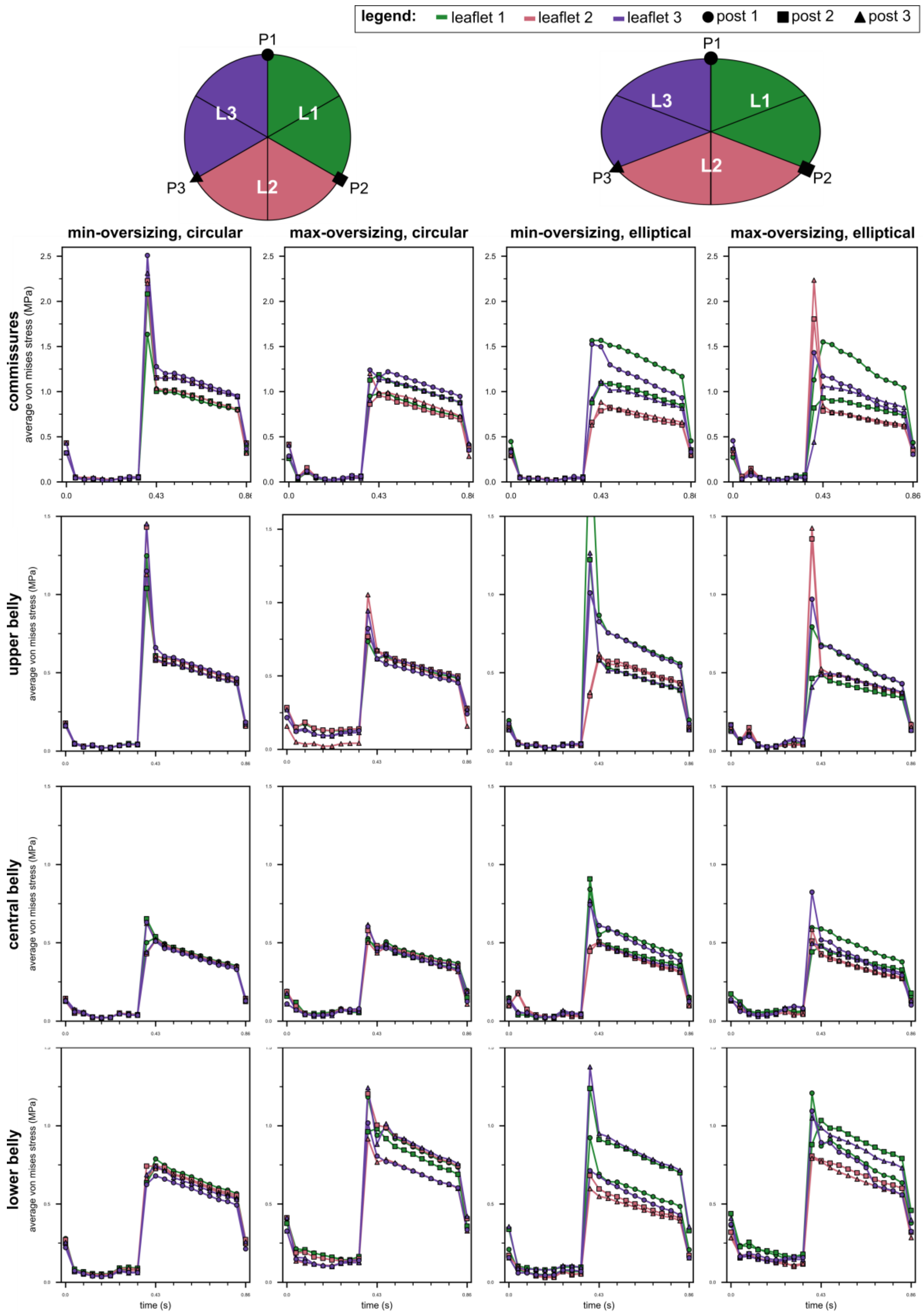


Figure S2: von Mises stress throughout the cardiac cycle for regions across bioprosthetic leaflet

*Table S1: Variation in the von Mises stress at peak diastole (t = 0.430 s)*

| **Region** | **Leaflet-Post** | **Min-oversizing, circular** | **Max-oversizing, circular** | **Min-oversizing, elliptical** | **Max-oversizing, elliptical** |
|---|---|---|---|---|---|
| Commissures | L1-P1 | 1.030 | 0.979 | 1.568 | 1.551 |
| | L1-P2 | 1.003 | 1.187 | 1.091 | 0.930 |
| | **Leaflet 1** | **1.016** | **1.083** | **1.330** | **1.240** |
| | L2-P2 | 1.153 | 0.960 | 0.788 | 0.785 |
| | L2-P3 | 1.036 | 0.980 | 0.884 | 0.843 |
| | **Leaflet 2** | **1.094** | **0.970** | **0.836** | **0.814** |
| | L3-P3 | 1.154 | 1.133 | 1.112 | 1.061 |
| | L3-P1 | 1.277 | 1.153 | 1.498 | 1.171 |
| | **Leaflet 3** | **1.215** | **1.143** | **1.305** | **1.116** |
| | **Mean** | **1.109** | **1.065** | **1.157** | **1.057** |
| | **STD** | **0.105** | **0.103** | **0.317** | **0.280** |
| Upper Belly | L1-P1 | 0.575 | 0.495 | 0.845 | 0.663 |
| | L1-P2 | 0.594 | 0.553 | 0.564 | 0.479 |
| | **Leaflet 1** | **0.584** | **0.524** | **0.704** | **0.571** |
| | L2-P2 | 0.567 | 0.566 | 0.584 | 0.474 |
| | L2-P3 | 0.594 | 0.548 | 0.609 | 0.514 |
| | **Leaflet 2** | **0.581** | **0.557** | **0.597** | **0.494** |
| | L3-P3 | 0.573 | 0.553 | 0.569 | 0.475 |
| | L3-P1 | 0.644 | 0.638 | 0.807 | 0.650 |
| | **Leaflet 3** | **0.609** | **0.596** | **0.688** | **0.563** |
| | **Mean** | **0.591** | **0.559** | **0.663** | **0.543** |
| | **STD** | **0.028** | **0.046** | **0.128** | **0.089** |
| Central Belly | L1-P1 | 0.528 | 0.449 | 0.501 | 0.579 |
| | L1-P2 | 0.533 | 0.472 | 0.547 | 0.473 |
| | **Leaflet 1** | **0.531** | **0.461** | **0.524** | **0.526** |
| | L2-P2 | 0.508 | 0.476 | 0.499 | 0.425 |
| | L2-P3 | 0.520 | 0.430 | 0.499 | 0.417 |
| | **Leaflet 2** | **0.514** | **0.453** | **0.499** | **0.421** |
| | L3-P3 | 0.512 | 0.445 | 0.483 | 0.474 |
| | L3-P1 | 0.505 | 0.460 | 0.604 | 0.511 |
| | **Leaflet 3** | **0.509** | **0.453** | **0.544** | **0.493** |
| | **Mean** | **0.518** | **0.456** | **0.522** | **0.480** |
| | **STD** | **0.011** | **0.017** | **0.046** | **0.060** |
| Lower Belly | L1-P1 | 0.727 | 0.881 | 0.635 | 0.819 |
| | L1-P2 | 0.678 | 0.920 | 0.850 | 0.974 |
| | **Leaflet 1** | **0.702** | **0.901** | **0.743** | **0.897** |
| | L2-P2 | 0.682 | 0.943 | 0.548 | 0.722 |
| | L2-P3 | 0.671 | 0.710 | 0.502 | 0.719 |
| | **Leaflet 2** | **0.677** | **0.826** | **0.525** | **0.721** |
| | L3-P3 | 0.687 | 0.824 | 0.881 | 0.924 |
| | L3-P1 | 0.631 | 0.750 | 0.627 | 0.841 |
| | **Leaflet 3** | **0.659** | **0.787** | **0.754** | **0.882** |
| | **Mean** | **0.679** | **0.838** | **0.674** | **0.833** |
| | **STD** | **0.031** | **0.094** | **0.157** | **0.104** |
| Attachment Edge | L1-P1 | 0.267 | 0.399 | 0.247 | 0.459 |
| | L1-P2 | 0.232 | 0.427 | 0.297 | 0.454 |
| | **Leaflet 1** | **0.249** | **0.413** | **0.272** | **0.457** |
| | L2-P2 | 0.270 | 0.422 | 0.255 | 0.431 |
| | L2-P3 | 0.250 | 0.323 | 0.233 | 0.396 |
| | **Leaflet 2** | **0.260** | **0.373** | **0.244** | **0.414** |
| | L3-P3 | 0.264 | 0.386 | 0.218 | 0.396 |
| | L3-P1 | 0.267 | 0.348 | 0.200 | 0.355 |
| | **Leaflet 3** | **0.265** | **0.367** | **0.209** | **0.376** |
| | **Mean** | **0.258** | **0.384** | **0.242** | **0.415** |
| | **STD** | **0.015** | **0.042** | **0.033** | **0.040** |

#### 2 Wall shear stress

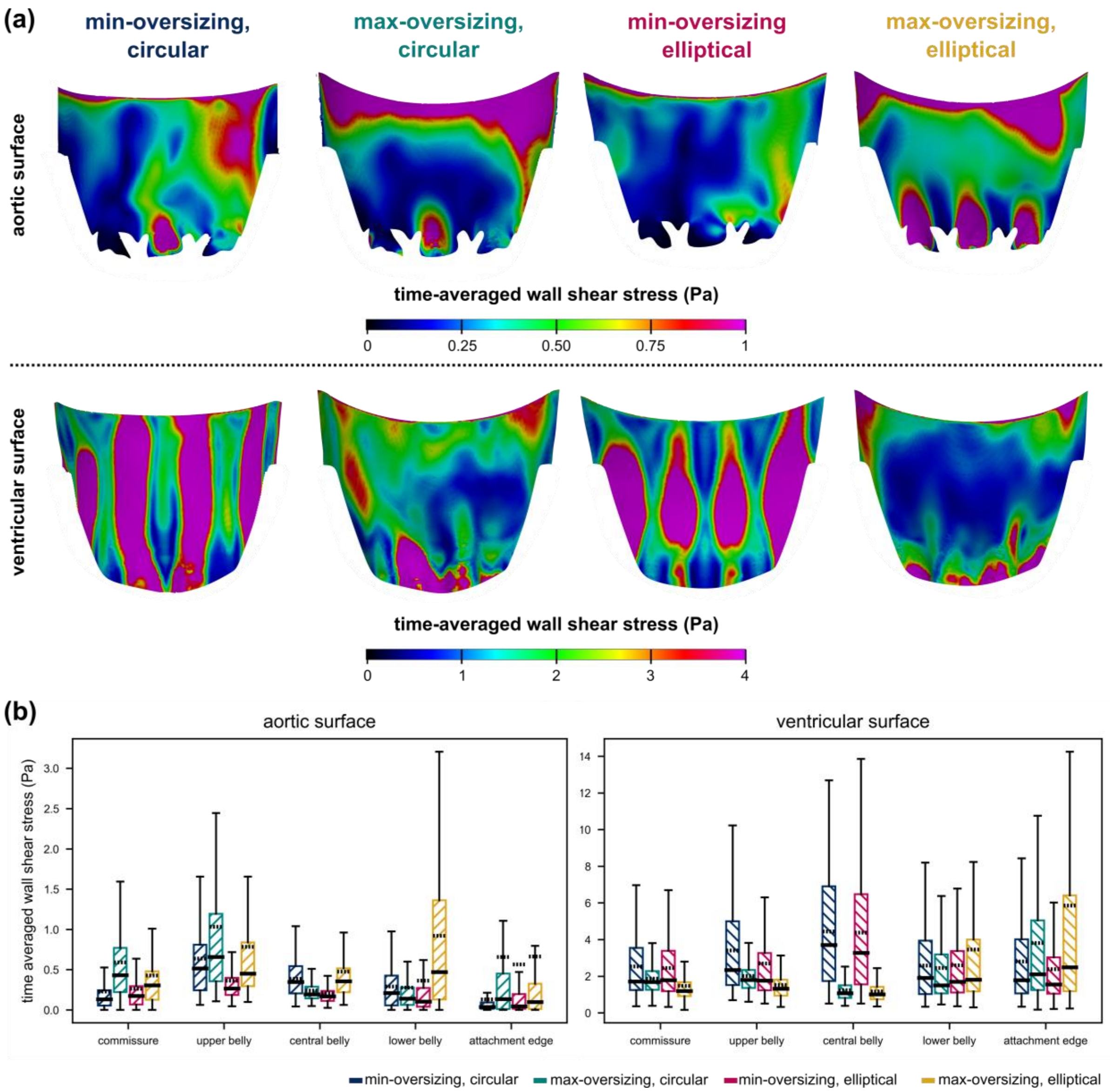


*Figure S3:(a) Contour plot of time-averaged wall shear stress (TAWSS). (b) TAWSS distribution.*

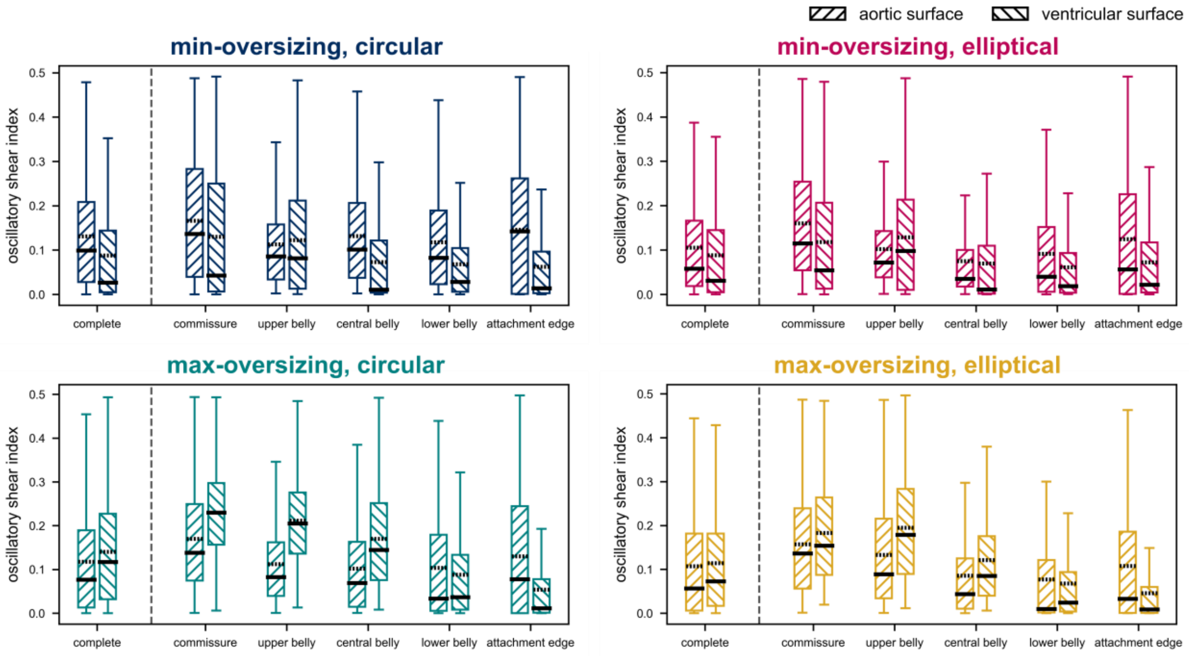


*Figure S4: Oscillatory shear index (OSI) distribution*

### 3 Stent Fatigue

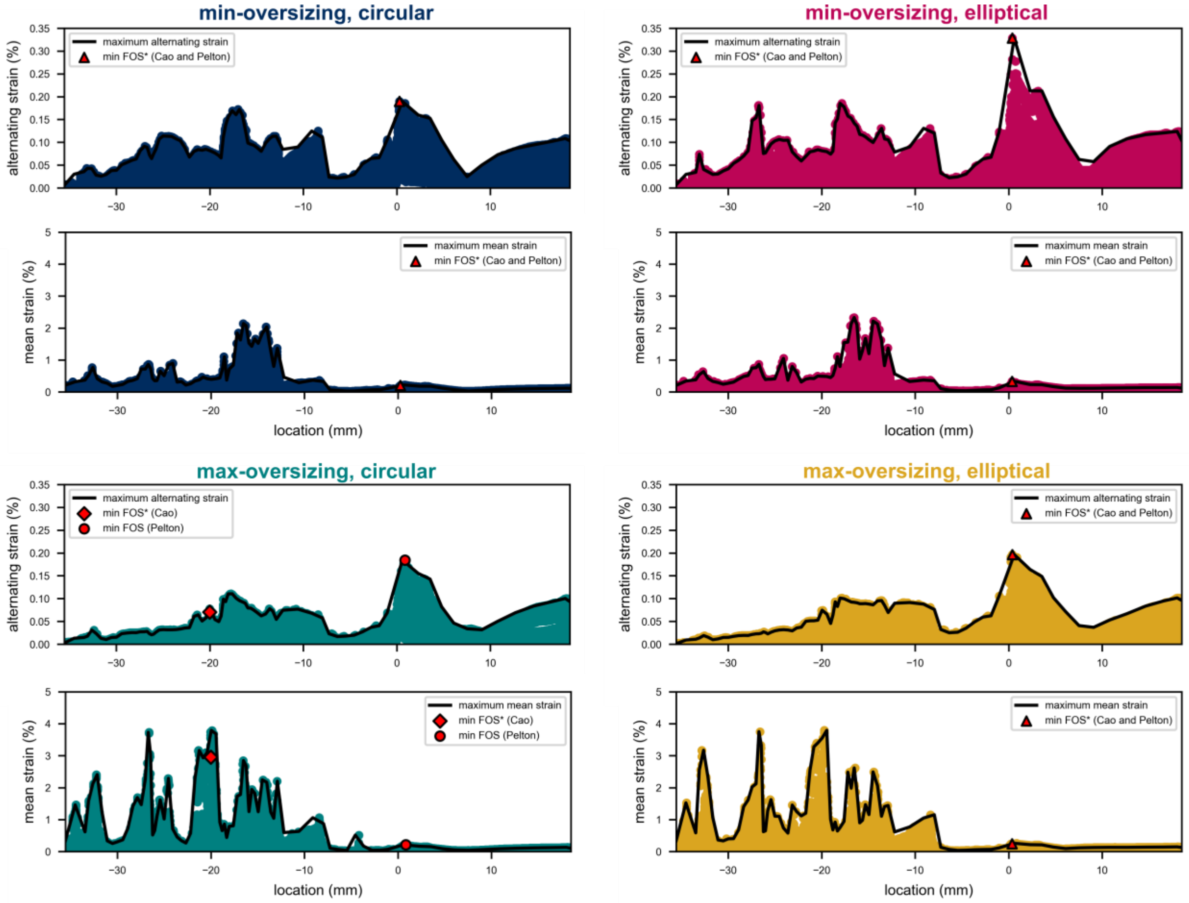


*Figure S5: Extraction of the maximum mean and alternating strain along the stent (black line) for all data points (coloured marker), also showing minimum fatigue FOS as per criteria proposed by Cao et al. (2017) and Pelton et al. (2008).*
